# Genetic variants predisposing to increased risk of kidney stone disease

**DOI:** 10.1101/2024.06.07.24308490

**Authors:** Catherine E. Lovegrove, Michelle Goldsworthy, Jeremy Haley, Smelser Diane, Caroline Gorvin, Fadil M. Hannan, Anubha Mahajan, Suri Mohnish, Omid Sadeghi-Alavijeh, Shabbir Moochhala, Daniel Gale, David Carey, Michael V. Holmes, Dominic Furniss, Rajesh V. Thakker, Sarah A. Howles

## Abstract

Kidney stones (KS) are common, heritable, and associated with mineral metabolism abnormalities. We used Mendelian randomization and colocalization to identify variants predicted to increase KS risk via increased serum calcium or decreased serum phosphate (odds ratios for genomic regions=4.30-13.83 per 1 standard deviation alteration) that account for 11-19% of KS due to reduced calcium-sensing receptor (CaSR)-signal transduction, increased urinary phosphate excretion, and impaired 1,25-dihydroxyvitamin D inactivation via diacylglycerol kinase delta (*DGKD*), solute carrier family 34 member 1 (*SLC34A1*), and cytochrome P450 family 24 subfamily A member 1 (*CYP24A1*), respectively. In silico analyses revealed that targeting *CASR*, *DGKD*, or *CYP24A1* to decrease serum calcium, or *SLC34A1* to increase serum phosphate may reduce KS risk, and in vitro studies demonstrated that positive CaSR-allosteric modulation ameliorates CaSR-signal transduction impaired by reduced DGKδ expression or KS-associated *DGKD* missense variants. These studies suggest that genotyping individuals with KS may facilitate personalized risk stratification and pharmacomodulation.

## Introduction

Kidney stone disease has a lifetime prevalence of ∼20% in men and ∼10% in women and is commonly a recurrent condition^1,2^. Systemic disorders, including disturbances of calcium homeostasis, may predispose to kidney stone formation and rare monogenic causes of nephrolithiasis are well recognised^3^. However, most cases of kidney stone disease are considered idiopathic and multiple genetic and environmental factors contribute to the observed phenotype^4,5^. The homeostatic and renal tubular mechanisms underlying these common forms of kidney stone disease are poorly understood, hampering efforts to implement improved therapeutic strategies to prevent recurrent kidney stone formation^6^. Genomic studies have revealed that higher serum calcium concentrations and lower serum phosphate concentrations likely increase risk of kidney stone disease^7^, suggesting that minor perturbations in mineral metabolism within the normal range may be a common risk factor for kidney stone formation. To further characterize the mechanisms by which alterations in calcium and phosphate homeostasis contribute to kidney stone disease we pursued genetic discovery studies, *in vitro* assays, and 3D-modelling to report kidney stone disease-causing variants associated with diacylglycerol kinase delta, *DGKD,* which encodes the calcium-sensing receptor (CaSR)-signaling partner DGKδ; solute carrier family 34 member 1, *SLC34A1,* which encodes the renal sodium-phosphate transport protein 2A, NaPi-IIa; and cytochrome P450 family subfamily A member 1, *CYP24A1,* which encodes 24-hydroxylase that inactivates 1,25-dihydroxyvitamin D.

## Results

### Putative kidney stone causal variants

To facilitate genetic analyses, a genome-wide association study (GWAS) was undertaken in the UK Biobank, considering 11,186 kidney stone disease cases and 390,488 controls. This GWAS identified 18 independent genetic signals at 17 loci (*ALPL, DGKD, SLC34A1, FLOT1, KCNK5, SLC22A2, HIBADH, TRPV5, AMPD3, DGKH, SEMA6D, ABCC6, UMOD, SOX9, CYP24A1, CLDN14,* and *GNAZ,* Figure 1, Figure S1-3, Table S1-2); associations at three of these loci (*FLOT1, SEMA6D, ABCC6*) have not previously been reported in kidney stone disease.

**Figure 1:**
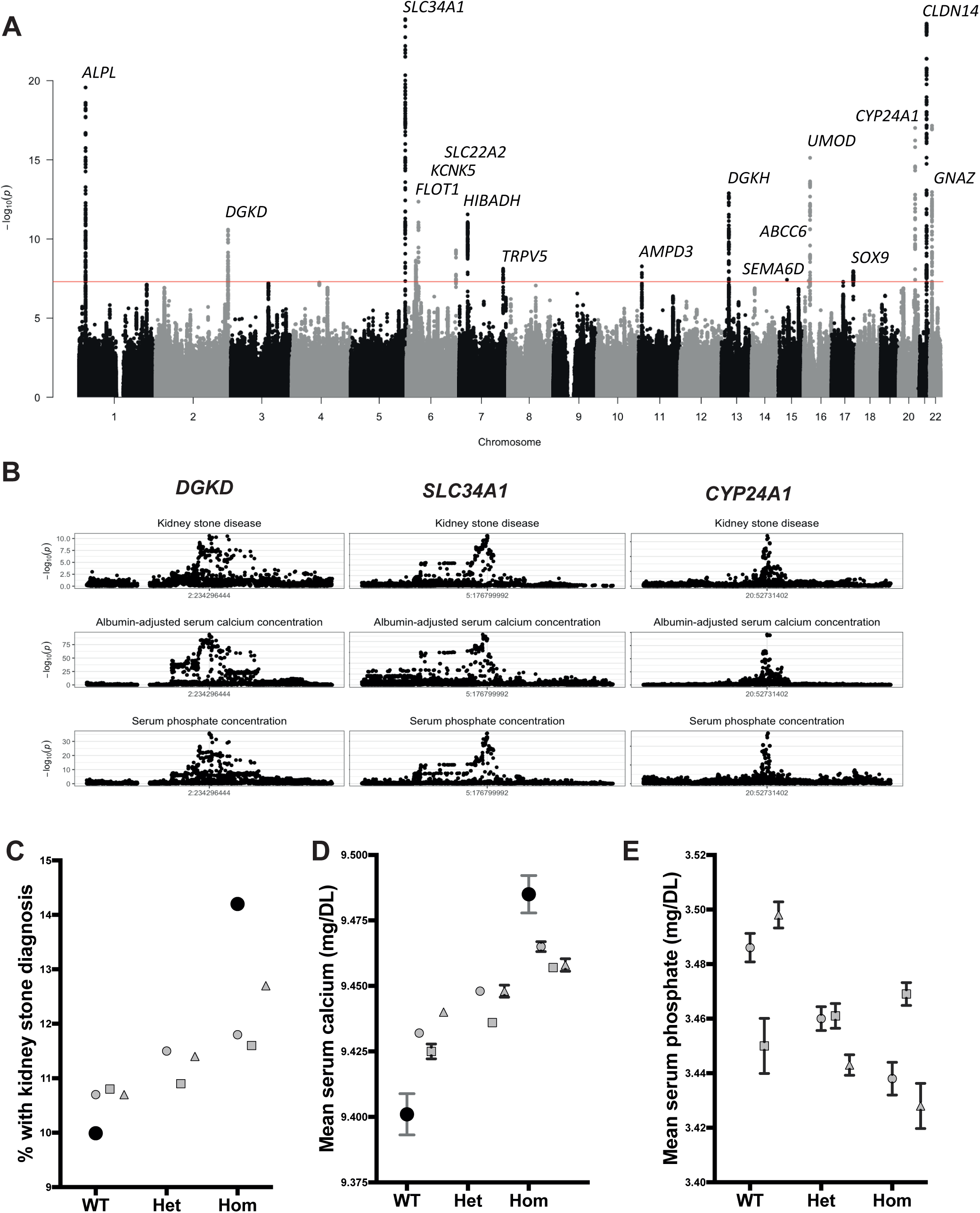
Genetic associations of kidney stone disease, serum calcium and phosphate concentrations. **A** Results of genome-wide association study (GWAS) of 11,186 kidney stone disease cases and 390,488 controls in the UK Biobank. Manhattan plot showing genome-wide p-values (-log10) plotted against their respective positions on each chromosome. The horizontal red line indicates the genome-wide significance threshold (5.0LxL10^−8^). Loci are labelled with the primary candidate gene at each locus. These loci are in proximity to biomineralization associated alkaline phosphatase, *ALPL*; diacylglycerol kinase delta, *DGKD;* solute carrier family 34 member 1, *SLC34A1*; flotillin 1, *FLOT1*; potassium two pore domain channel subfamily K member 5, *KCNK5*; solute carrier family 22 member 2, *SLC22A2;* 3-hydroxyisobutyrate dehydrogenase, *HIBADH*; transient receptor potential cation channel subfamily V member 5, *TRPV5*; adenosine monophosphate deaminase 3, *AMPD3*; diacylglycerol kinase eta, *DGKH*; semaphoring 6D, *SEMA6D*; ATP binding cassette subfamily C member 6, *ABCC6*; uromodulin, *UMOD*; SRY-box transcription factor, *SOX9*; cytochrome P450 family 24 subfamily A member 1, *CYP24A1*; claudin 14, *CLDN14*; and G protein subunit alpha z, *GNAZ*. Three loci (*FLOT1, ABCC6,* and *SEMA6D*) have not been previously associated with kidney stone disease. **B** Locus zooms from GWAS studies of kidney stone disease, albumin-adjusted serum calcium, and serum phosphate concentrations in the UK Biobank at loci where there is evidence from regional MR that risk of kidney stone disease is increased via effects on serum calcium and phosphate homeostasis and where genetic associations of kidney stone disease, serum calcium and phosphate concentrations colocalize. **C-E** Associations of genotype with kidney stone disease (**C**), serum calcium concentration (**D**), and serum phosphate concentration (**E**) in the DiscovEHR cohort (11,451 kidney stone cases; 86,294 controls). Mean serum calcium (**D**) and phosphate (**E**) measurements ±standard error of the mean (SEM) are adjusted for kidney stone disease case status. Note, in panel D, the SEM is very small for most concentrations and obscured by the graphical icon. Associations of combinations of *DGKD-, CYP24A1-,* and *SLC34A1-* risk alleles were not assessed for serum phosphate due to lack of directional concordance. These findings provide evidence to support the variants rs838717, rs10051765, and rs6127099 as causal risk factors for kidney stone disease acting via reduced CaSR signal transduction, impaired vitamin-D inactivation, and increased urinary phosphate excretion, respectively.

To ascertain regional effects of genetically-predicted serum calcium and phosphate concentrations on odds of kidney stone disease, a cis-Mendelian randomization approach was employed, systematically considering 1MBp genomic areas ±500kbp of lead independent variants associated with serum albumin-adjusted calcium or phosphate at GWAS (Supplementary appendix). Where potential causal regional effects of serum calcium or phosphate concentrations on kidney stone disease were identified, colocalization analyses were used to evaluate the probability of a single shared causal variant^8,9^ considering data from kidney stone disease, serum albumin-adjusted calcium, phosphate, and parathyroid hormone (PTH) concentrations GWAS simultaneously (Figure S1)^10–12^ . Three variants were identified that were significantly associated with kidney stones and predicted to causally increase kidney stone disease risk via effects on serum calcium and phosphate homeostasis (Figure 1, Figures S1 and S4, Tables S3-S5, Supplementary data 2-3). These variants comprised: an intronic *DGKD* variant that is a predicted transcription factor binding site (rs838717) with a MR estimate (odds ratio, OR) of regional effects of a 1 standard deviation (SD, 0.08mmol/L) increase in albumin-adjusted serum calcium on kidney stone disease of 4.30 (95% confidence interval (CI)=2.81-6.58, posterior probability SNP is causal variant (PP)=1.00); an intergenic variant ∼6kb upstream of *SLC34A1* (rs10051765) with a MR estimate of regional effects of a 1SD (0.16mmol/L) decrease in serum phosphate on kidney stone disease of 13.83 (95% CI=9.44-20.27, PP=0.97); and an intergenic variant ∼50kb downstream of *CYP24A1* (rs6127099) with a MR estimate of regional effects of a 1SD increase in albumin-adjusted serum calcium on kidney stone disease of 11.42 (95% CI=8.41-15.5, PP=1.00) (Tables S3-S6).

Mutations of *CYP24A1* and *SLC34A1* are known to cause infantile hypercalcaemia (IH) types 1 and 2, respectively^13,14^, which are autosomal recessive disorders of calcium and phosphate metabolism associated with nephrocalcinosis and kidney stone disease. IH1 is due to impaired inactivation of 1,25-dihydroxyvitamin D, which leads to elevations in circulating 1,25-dihydroxyvitamin D that results in increased intestinal and renal absorption of calcium with consequent hypercalcaemia. In contrast, IH2 is caused by increased renal phosphate excretion due to impaired NaPi-IIa function resulting in a reduction in serum fibroblast growth factor 23 concentrations, activation of 1-α hydroxylase (an enzyme that activates 25-hydroxyvitamin D) and inhibition of 24-hydroxylase. In addition, a reduction in DGKδ expression results in impaired CaSR-signal transduction^15^, and gain- and loss-of-function mutations in components of the CaSR-signaling pathway cause autosomal dominant hypocalcaemia (ADH) with relative hypercalciuria and familial hypocalciuric hypercalcaemia (FHH), respectively^16,17^. We sought associations of genotype with serum biochemistry in the DiscovEHR cohort and using UK Biobank GWAS data (Supplementary appendix). Our results reveal that the predicted *DGKD* (rs838717) and *SLC34A1* (rs10051765) causal variants are associated with higher serum calcium and lower serum phosphate concentrations which are consistent with attenuated forms of FHH and IH2, respectively, and that the predicted *CYP24A1* (rs6127099) causal variant is associated with higher serum calcium and phosphate concentrations, consistent with an attenuated form of IH1 (Figure 1, Table S7-8).

To determine the clinical relevance of these variants in conferring risk of developing kidney stone disease, we calculated the fraction of kidney stone disease that may arise due to these three putative causal variants. This revealed a population attributable fraction of ∼11% in DiscovEHR and ∼19% in UK Biobank (Tables S9-10). Furthermore, addition of a single *DGKD-*, *SLC34A1-,* or *CYP24A1-,* putative causal variant was associated with a 6-10%, 10-16%, and 5-14%, increased odds of kidney stone disease, respectively, and occurrence of all 6 risk alleles in an individual was associated with a ∼4% increased prevalence and >35% increased odds of kidney stone disease (Tables S7-8). Thus, *DGKD, SLC34A1,* and *CYP24A1* variants confer risks that summate to a substantially increased risk of developing kidney stone disease. Moreover, these findings indicate that reduced CaSR signal transduction, increased urinary phosphate excretion, and impaired vitamin D inactivation may be common causes of kidney stone disease.

### Drug-target Mendelian Randomization

To identify potential therapeutic pathways that could be modulated to prevent kidney stone disease, we undertook drug-target MR analyses using a stringent threshold of *r*^2^<0.01 to define independence of genetic variants as exposure instrumental variables. GWAS summary statistics from studies in the UK Biobank, and the FinnGen study were used as outcome datasets (Figures S1-2). These analyses suggested that modulating *CASR* and *CYP24A1* to reduce serum calcium concentrations by 1SD may decrease kidney stone disease relative risk by ∼30% and ∼90%, respectively (Figure 2, Table S11); directionally concordant but statistically insignificant effects were detected for *CASR*-mediated effects using FinnGen outcome data (Figure 2, Table S11). Similar analyses of *DGKD* or *SLC34A1* modulation were not possible as there were insufficient genetic proxies. However, MR analyses relaxing the threshold of genetic independence of instrumental variables to *r*^2^<0.1, indicated that reducing serum calcium concentrations by 1SD via *DGKD* may decrease risk of kidney stone disease by ∼70%, and increasing serum phosphate concentrations by 1SD via *SLC34A1* may decrease risk of kidney stone disease by >90% (Figure 2, Table S11). Phenome wide association study data suggested that modulating *DGKD, CASR, CYP24A1,* or *SLC34A1* may result in target-mediated adverse effects including alterations in serum bilirubin, inflammatory bowel disease, migraine, atopic dermatitis, and eczematous phenotypes (Supplementary data 4).

**Figure 2:**
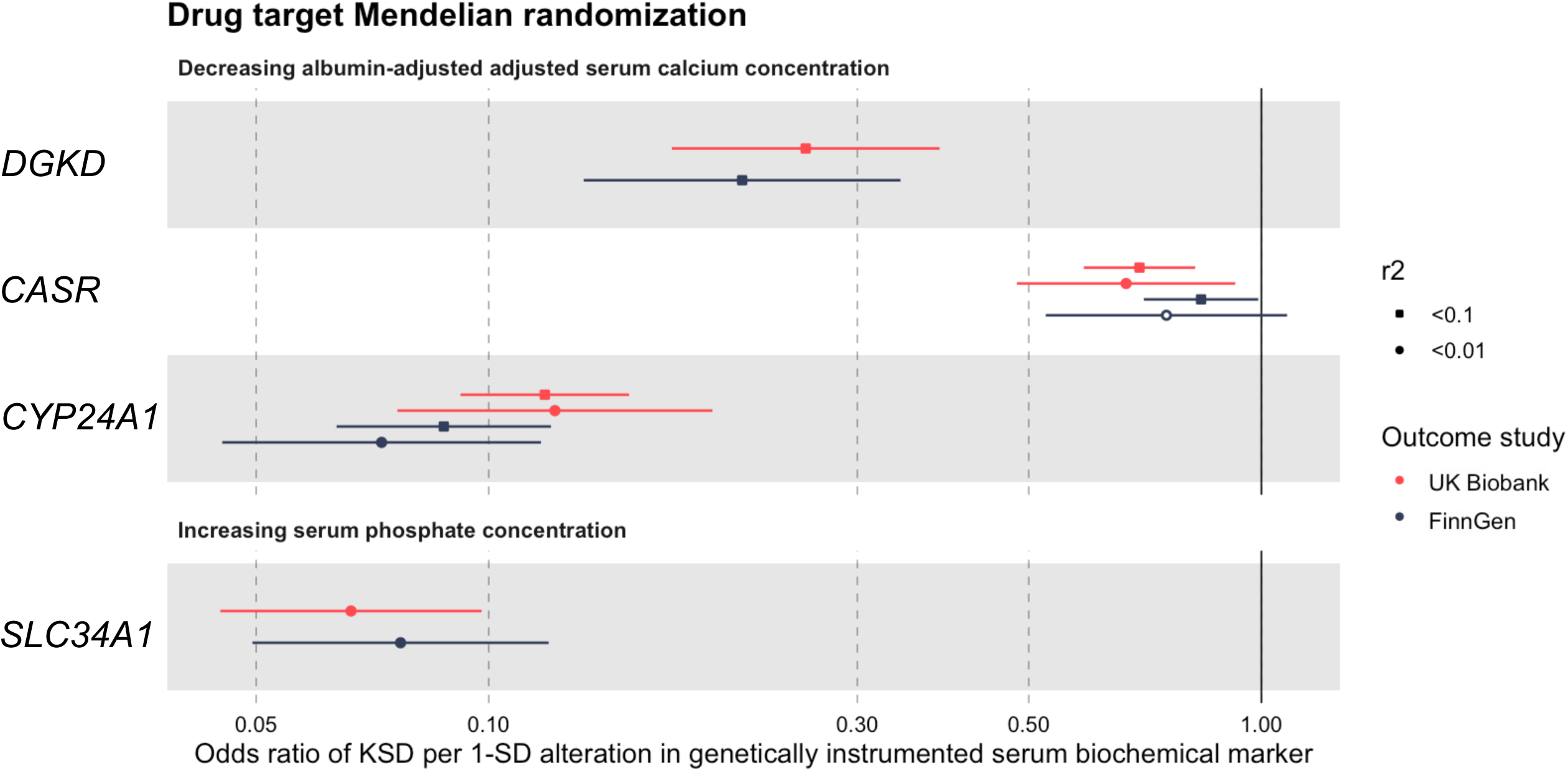
Drug target Mendelian randomization. Forest plot of predicted effects of modulating albumin-adjusted serum calcium concentrations via *DGKD, CASR,* or *CYP24A1* or serum phosphate concentrations via *SLC34A1.* KSD=kidney stone disease; SD=standard deviation. Gene positions are defined via Ensembl +/-300kbp. There were insufficient genetic instruments to undertake analyses of modulating serum calcium or phosphate concentrations via *DGKD* or *SLC34A1* using a threshold for genetic independence (r^2^) of 0.01. These data indicate that reducing serum calcium via *DGKD, CASR, CYP24A1,* or increasing serum phosphate via *SLC34A1* would decrease the risk of kidney stone disease.

### Coding region DGKD variants associated with kidney stone disease

The function of *SLC34A1* and *CYP24A1* in mineral metabolism and IH have been well characterized^13,18^ and we therefore focused on further defining the role of DGKδ in CaSR signaling and kidney stone disease. A total of 7 rare, predicted deleterious *DGKD* variants associated with kidney stone disease were identified in the Genomics England 100,000 genomes (100kGP) and DiscovEHR cohorts (2 from the 100kGP, His190Gln and Ile221Asn; 4 from DiscovEHR, Ile91Val, Thr319Ala, Val464Ile, Arg900His; and 1, Arg1181Trp, from both cohorts) (Tables S12-13). Residues Ile91, His190, Ile221, Thr319, Arg900, and Arg1181 are evolutionary conserved, and residue Val464 partially conserved, across vertebrate DGKδ orthologues suggesting that these DGKδ variants may be pathogenic (Figure S5). In DiscovEHR, 6 kindreds with *DGKD* variants comprised 13 individuals who were variant carriers and affected with a relevant phenotype (11 kidney stone disease, 1 hypercalciuria, and 1 primary hyperparathyroidism and kidney stone disease); 7 individuals who were variant carriers but unaffected; 12 individuals who were not variant carriers and unaffected; and 3 individuals who were not variant carriers but affected with kidney stone disease (n=2), or primary hyperparathyroidism (n=1) (Figure 3). Variants Thr319Ala and Val464Ile co-segregated with kidney stone disease in two DiscovEHR cohort kindreds but penetrance was incomplete for Val464Ile (Figure 3). Statistically significant associations of Arg1181Trp with kidney stone disease were not detected in DiscovEHR, however, in three kindreds Arg1181Trp co-segregated with kidney stone disease with incomplete penetrance, and in a further kindred, co-segregation was incomplete with the possibility of additional genetic risk factors for kidney stone disease (Figure 3).

**Figure 3:**
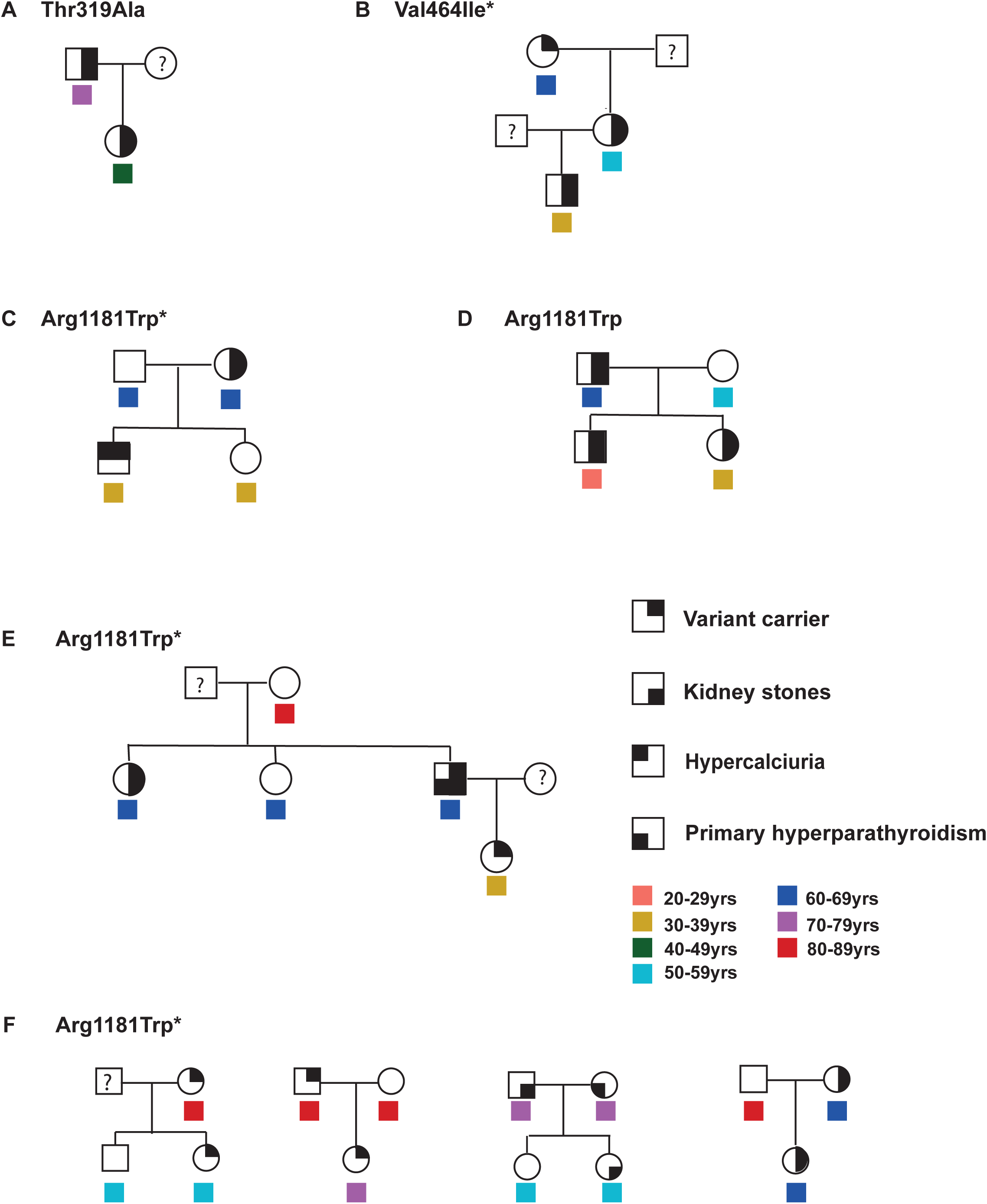
Family trees of DiscovEHR kindreds carrying *DGKD* variants. Squares represent male family members, circles female family members,? indicates missing data. Individuals ages are represented by coloured symbols. To protect the identity of kindreds, some information has been indicated by *. In the case of kindred F, simple pedigrees only are shown, full information can be provided by contacting the corresponding author.

### Functional characterization of DGK**δ** mutants

The effects of kidney stone disease-associated *DGKD*-variants and reduced DGKδ expression on CaSR-signal transduction were assessed in CaSR-expressing HEK293 cells and their responses to alterations in extracellular calcium concentration determined using SRE and NFAT-RE assays to evaluate signaling via Ras-Raf-MEK-ERK and intracellular calcium release, respectively (Figure S9). Variants Ile91Val, His190Gln, Ile221Asn, Thr319Ala, Val464Ile, Arg900His and Arg1181Gly resulted in reduced SRE-and/or NFAT-RE mediated responses in comparison to cells transfected with wild-type DGKδ; and reduced DGKδ expression, resulting from shRNA *DGKD* knockdown, attenuated SRE-mediated responses without change in NFAT-mediated responses (Figures 4 and S6-7). These findings are consistent with loss-of-function mutations in components of the CaSR-signaling pathway and indicate that DGKδ knockdown results in biased CaSR-signal transduction. The CaSR positive allosteric modulator, cinacalcet, rectified CaSR-signaling loss-of-function effects associated with reduced DGKδ expression and ameliorated impaired SRE-responses due to DGKδ kidney-stone mutations (Figure 4). However, cinacalcet had no effect on impaired NFAT-RE responses due to DGKδ kidney-stone mutations, except for the mildly inactivating Arg900His (Figures 4 and S7).

**Figure 4:**
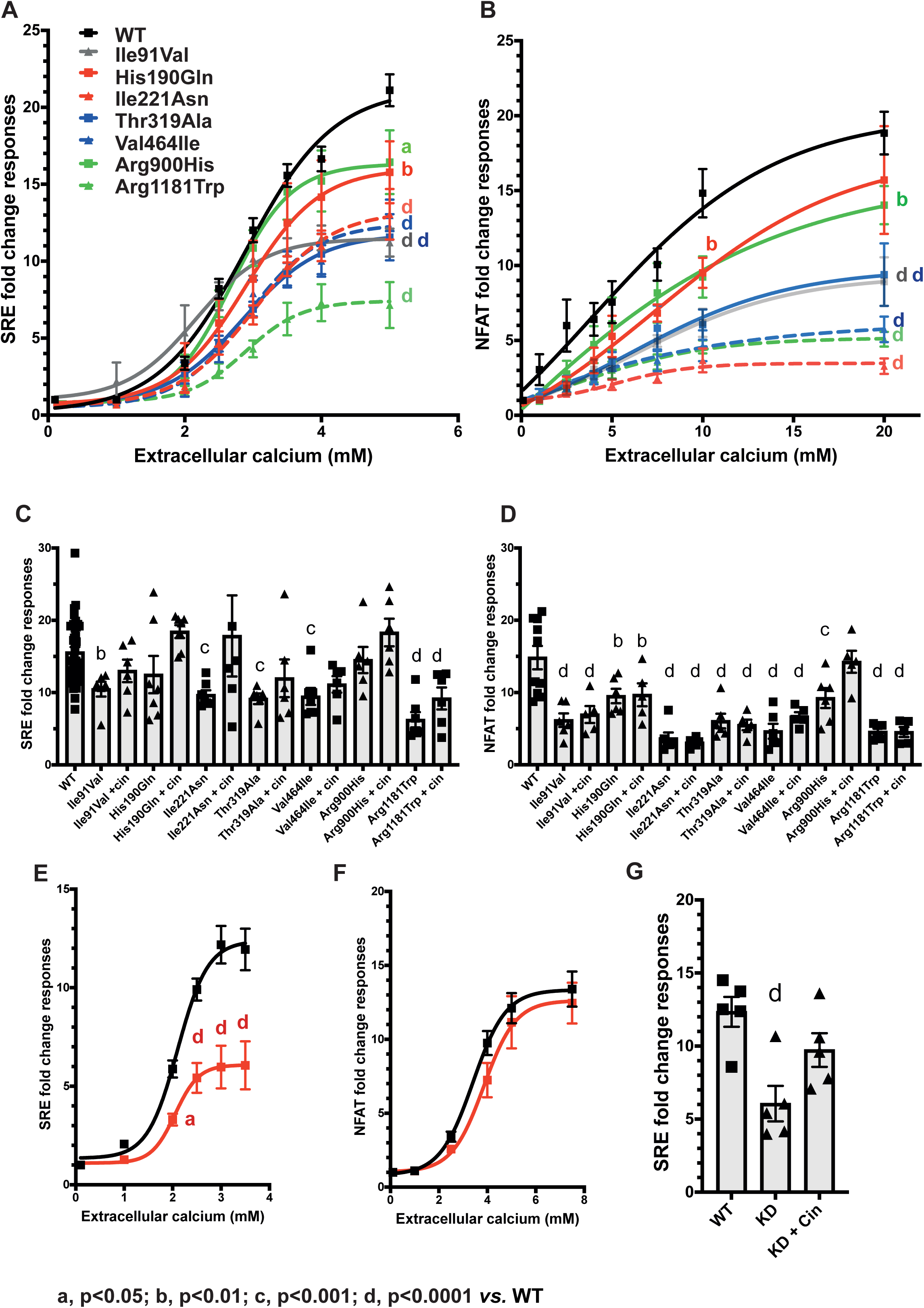
Functional characterization of kidney stone-associated DGKδ variants. (**A**) CaSR-mediated SRE and (**B**) NFAT-RE responses to changes in extracellular calcium concentration [Ca^2+^]_e_ in HEK-CaSR-DGKδ cells stably transfected with wild-type (WT) or kidney stone-associated variants Ile91Val, His190Gln, Ile221Asn, Thr319Ala, Val464Ile, Arg900His, and Arg1181Trp. Transfection with kidney stone-associated *DGKD* variants led to a reduction in SRE- and NFAT-RE-responses compared to cells transfected with wildtype (WT) *DGKD*. (**C**) Effect of 100nM cinacalcet (cin) treatment on SRE-responses at 3.5mM [Ca^2+^]_e_ and (**D**) NFAT-RE-responses at 10mM [Ca^2+^]_e_ in HEK-CaSR-DGKδ cells transfected with the kidney stone-associated variants. Treatment with cinacalcet increased SRE-mediated responses of all variants but had no effect on NFAT-RE responses except for cells transfected with the Arg900His variant. (**E**) CaSR-mediated SRE and (**F**) NFAT-RE responses to changes in extracellular calcium concentration [Ca^2+^]_e_ in HEK-CaSR cells following DGKδ knockdown; DGKδ knockdown (red) led to a reduction in SRE-responses without change in NFAT-RE-responses, compared to WT (black). (**G**) Effect of 5nM cinacalcet (cin) treatment on SRE-responses at 3.5mM [Ca^2+^]_e_ in HEK-CaSR cells following DGKδ knockdown. Treatment with cinacalcet rectified impaired SRE-mediated responses. Mean fold change responsesL± standard error of the meanL(SEM) are shown for n>4 biologically independent experiments. Two-way ANOVA with Dunnett’s correction for multiple comparisons was used to compare points on dose response curve with reference to wild-type. These data provide evidence that kidney stone disease is associated with impaired CaSR-signal transduction, which can be ameliorated with cinacalcet.

### Predicted effects of DGK**δ** variants on protein function

To further elucidate the mechanisms by which kidney stone disease associated DGKδ variants may alter DGKδ function we pursued three-dimensional modelling studies. DGKδ Arg1181 is in the sterile alpha motif (SAM) domain, which facilitates DGKδ oligomerization and intracellular localization^19^. Analysis of the crystal structure of oligomeric DGKδ SAM domains indicated that Arg1181 likely forms a polar contact with Asp1183 on adjacent DGKδ SAM domains. Replacing the polar Arg1181 residue with a nonpolar Trp1181 residue is predicted to cause a reduced affinity for the adjacent DGKδ SAM domain, which may compromise DGKδ oligomerization and alter intracellular localization (Figure 5). Mutation of Asp1183 to Gly1183 is reported to increase DGKδ solubility *in vitro*, indicating a reduction in oligomerization, and induce spontaneous localization of DGKδ to the plasma membrane^20^. As the crystal structure of the remainder of DGKδ has not been solved, we undertook additional analyses using the AlphaFold DGKδ predicted structure. This revealed that DGKδ Ile91 is within the pleckstrin homology domain and the Val91 variant may impair DGKδ binding to partner proteins and cell membrane localisation^21,22^; His190 and Ile221 are within the C1 domain of DGKδ, and the Gln190 and Asn221 variants may affect diacylglycerol (DAG) binding^23^; and Thr319 and Arg900 are located in proximity to the ATP-binding pocket and in the accessory domain ATP-binding motif, respectively, and the Ala319 and His900 variants may alter ATP-binding dynamics (Figure S8). Predictions regarding the mechanistic effects of Val464Ile could not be made due to its location in a region of the AlphaFold structure with very low model confidence.

**Figure 5:**
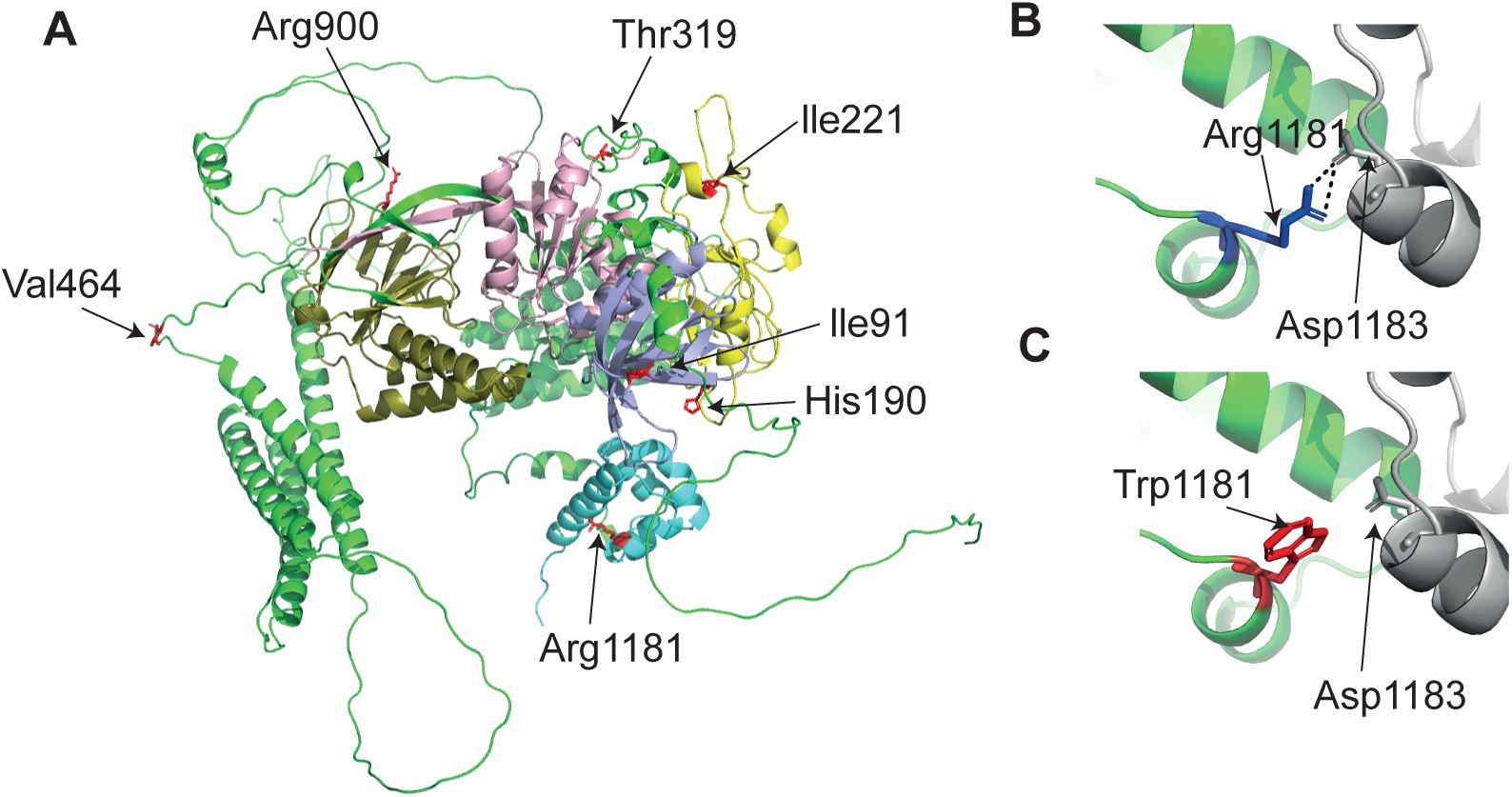
Three-dimensional modelling of kidney stone-associated DGKδ variants. **A:** Predicted structure of DGKδ isoform 2 ((AF-Q16760-F1-mod^45,46^, AlphaFold). Residue Ile91 lies in the pleckstrin homology domain (purple); residues His190 and Ile221 are located in the cysteine rich domain (yellow); residue Thr319 is in the catalytic domain (pink); residue Val464 is in a linker region; residue Arg900 is in the accessory catalytic domain (dark green); and Arg1181 is located in the sterile alpha motif (SAM) domain (blue)^51^. **B:** Location of Arg1181 (dark blue) in oligomeric DGKδ SAM domain crystal structure (PDB 3BQ7^20^). Arg1181 is predicted to form a polar contact (dashed black line) with Asp1183 on adjacent DGKδ SAM structure (grey). **C:** Location of Trp1181 (red) in oligomeric DGKδ SAM domain crystal structure. Trp1181 is not predicted to form a polar contact with Asp1183 on adjacent DGKδ SAM structure (grey).

## Discussion

To our knowledge, this study is the first to employ a systematic, region-specific, MR approach, combined with colocalization analyses, to identify putative disease-causing genetic variants and pathways. Using this approach, we identified three common non-coding variants that predict an increased risk of kidney stone disease via DGKδ-mediated reduced CaSR-signal transduction, NaPi-IIa-mediated increased renal phosphate excretion, and 24-hydroxylase-mediated decreased 1,25-dihydroxyvitamin D inactivation. Furthermore, we find that these three variants may be responsible for 11-19% of kidney stone disease cases and that homozygosity for all putative causal variants is associated with a >35% increased odds and ∼4% increased prevalence of kidney stones.

These results may have direct clinical utility in facilitating prediction of kidney stone recurrence risk and thereby motivate lifestyle modifications and selection of therapeutic interventions, such as calcimimetics to ameliorate DGKδ-mediated CaSR-signaling perturbations; phosphate supplements to increase serum phosphate in NaPi-IIa-associated kidney stone disease; or inhibitors of vitamin D activation (e.g.: triazole drugs or rifampicin) and/or avoidance of vitamin D supplementation where 24-hydroxylase-mediated 1,25-dihydroxyvitamin D inactivation is likely impaired. Indeed, our drug target-MR studies indicate that modulating DGKδ, CaSR, or 24-hydroxylase to decrease serum calcium or NaPi-IIa to increase serum phosphate is predicted to decrease kidney stone disease risk by up to 90%. Furthermore, we demonstrate that cinacalcet, a positive CaSR allosteric modulator, ameliorates impaired CaSR-mediated signaling due to rare kidney stone associated-DGKδ missense mutations and normalizes biased CaSR-signal transduction in DGKδ-deplete cells. Idiopathic hypercalciuria occurs in up to 50% of individuals with kidney stone disease^24^ and negative CaSR-allosteric modulators have been suggested as a potential therapy to prevent kidney stones in these patients via effects on urinary calcium excretion^25^; our studies indicate this approach may actually increase incidence of kidney stone disease via effects on serum calcium, and that the role of increased serum calcium concentrations independent of urinary calcium concentrations in kidney stone risk has been underappreciated.

Idiopathic calcium oxalate stones may form on subepithelial renal papilla calcium phosphate aggregates known as Randall’s plaques. Randall’s plaques are hypothesised to arise due to high levels of calcium reabsorption in the thick ascending limb of the loop of Henle leading to increased calcium loads in the descending vasa recta which raise supersaturations sufficiently to cause mineral precipitation^26^. A tendency to higher serum and urinary calcium concentrations would act in tandem to increase the likelihood of plaque formation and may provide mechanistic insight into the occurrence Randall’s plaques and kidney stones in the absence of overt hypercalcemia or hypercalciuria.

Both FHH and ADH have been reported to be associated with kidney stone disease^27–30^, although kidney stones are more commonly occur in the context of ADH with hypercalciuria. Our current study reveals pathways linking the *DGKD*-associated predicted transcription factor binding site, rs838717, to kidney stone disease via increased serum calcium concentrations, a phenotype in keeping with decreased CaSR-signal transduction and FHH. Moreover, our functional studies of rare, coding kidney stone disease-associated *DGKD* variants are consistent with DGKδ-mediated impaired CaSR-signal transduction in the pathophysiology of nephrolithiasis. Kidney stones, parathyroid hyperplasia, mild hypercalcaemia, and hypercalciuria have been described in a kindred with the FHH-associated CaSR loss-of-function mutation Phe881Leu^27^. This is similar to our findings of hyperparathyroidism in individuals with kidney stone disease carrying DGKδ Gln190 and Trp1181 variants. Furthermore, DGKδ Trp1181 was associated with hypercalciuria in this study and rs838717 has been reported to be associated with higher urinary calcium excretion in a small cohort^15^, although associations of 24-hour urinary calcium excretion and rs838717 have not been replicated in a GWAS of ∼6,500 individuals^31^. Thus, DGKδ-associated perturbations of CaSR-signal transduction may have intracellular and tissue-specific effects that mirror CaSR loss-of-function variants associated with hyperparathyroidism phenotypes^27,32^.

Our findings of impaired DGKδ-mediated CaSR-NFAT-RE responses which are not ameliorated by cinacalcet and arise due to DGKδ missense variants but not DGKδ knockdown, provides evidence for DAG and inositol 1,4,5-trisphosphate cross-talk, and may indicate that DGKδ missense variants can exert dominant negative effects^33^. Furthermore, our studies indicate the importance of DGKδ oligomerization, ATP-binding, pleckstrin homology and cysteine-rich domains in DGKδ function. Monoallelic *SLC34A3* variants likely increase kidney stone risk via effects that are insufficient to cause fully penetrant Mendelian disease but confer a higher risk than aggregate effects of known common risk alleles^34^. The identification of incompletely penetrant DGKδ variants in this study, including the recurrent Arg1181Trp which is reported four times in a homozygous state in Gnomad, suggests that *DGKD* variants may fall into a comparable intermediate-risk category.

In conclusion, this study defines putative kidney stone disease-causing variants and demonstrates the central role of DGKδ-mediated reduced CaSR-signal transduction, increased renal phosphate excretion, and perturbed 1,25 vitamin D inactivation in kidney stone pathogenesis. Furthermore, our studies reveal potential novel therapeutic approaches for kidney stone prophylaxis and may have clinical utility in enabling personalized risk stratification and management strategies in kidney stone disease.

## Methods

### Ethical approval

UK Biobank has approval from the North West Multi-Centre Research Ethics Committee (11/NW/0382). Ethical approval for the 100,000 genomes (100KGP) was granted by the Cambridge South Research Ethics Committee for the East of England (REC Ref14/EE/1112). Additional informed consent was obtained from 100KGP participants using protocols approved by Multicenter Research Ethics Committee (MREC/02/2/93). Participants from the DiscovEHR cohort provided written informed consent for participation in the MyCode Community Health Initiative, an institutional review board–approved project (Protocol 2006-0258) that allows for genetic analysis and linking to information from the electronic health records. The research included in this publication was reviewed and determined to be exempt by the Geisinger IRB, #2023-1786. All participants gave written informed consent.

### Genome-wide association study (GWAS), Mendelian randomization (MR), and colocalization

A GWAS of kidney stone disease, including genetic sex and genotyping platform as covariates, was undertaken of the UK Biobank data^35^ using a linear mixed non-infinitesimal model (BOLT-LMM v2.4) LMM to account for population sub-structure and cryptic relatedness^36^ (Supplementary appendix). Regional (1MBp) effects of genetically-predicted serum calcium and phosphate concentrations on odds of kidney stone disease were estimated (TwoSample MR package, Rv4.3.1^37^), by selecting independent (r^2^<0.1) genetic variants ±500kbp of lead independent variants from serum albumin-adjusted calcium or phosphate GWAS significantly (p<5×10^-8^) associated with biochemical traits for use as instrumental variables^10^. Primary MR analyses used the inverse-variance weighted method and individual MR estimates were calculated using the Wald ratio. MR-Egger intercept and estimate were used to explore pleiotropic relationships; where the MR-Egger intercept was significantly different from zero (p<0.05) the MR-Egger estimate was interpreted as the estimate of best fit. Results are presented as effect estimates and corresponding 95% confidence intervals per standard deviation decrease in mineral metabolism trait on odds of KSD. Cochrane’s Q test was used to identify heterogeneity in causal estimates. P-values for MR estimates were adjusted for multiple testing using the Benjamini-Hochberg false discovery rate method, controlled at 5%^38^.

### Colocalization analyses

Once potential causal regional effects of serum calcium or phosphate concentrations on kidney stone disease were identified, colocalization analyses (*Coloc()* and HyPrColoc, Rv4.3.1) were used to evaluate the probability of a single shared causal variant^8,9^ and identify putative casual variants, considering data from kidney stone disease, serum albumin-adjusted calcium, phosphate, and parathyroid hormone (PTH) concentrations GWAS (Figure S1)^10–12^. *Coloc()* integrates evidence over all variants at a locus to enable one to evaluate the following hypotheses^8,9^: H0: Genomic region not associated with kidney stone disease or mineral metabolite trait; H1: Genomic region associated with mineral metabolite trait but not kidney stone disease; H2: Genomic region associated with kidney stone disease but not mineral metabolite trait; H3: Genomic regions associated with mineral metabolite trait and kidney stone disease with two separate putative causal variants; H4: Genomic region associated with mineral metabolite trait and kidney stone disease with one putative causal variant. coloc.abf() in colocR package with prior probabilities set to p1 = 1×10^-4^, p2 = 10×10^-4^, and p12 = 1×10^-5^ was used and results with PP H4 >0.75 were considered to show strong evidence of colocalization^8^.

### Drug target Mendelian randomization and phenome-wide association studies

The potential utility of modulating drug targets to prevent kidney stone disease was estimated using genetic proxies ±300kbp of target genes significantly associated (p<5×10^-8^) with relevant mineral metabolism traits (Figure S1)^10^. Using smaller genomic regions defined by target-gene coordinates, rather than 1Mbp loci used during regional MR, provided enhanced estimates of clinical utility of target modulation (Figure S2). To externally validate our findings we used publicly available FinnGen r8 GWAS data for the phenotype “N14 Calculus of kidney and ureter” (comprising 8,597 cases and 333,128 controls)^39,40^. MR analyses were performed using the same principles as described above.

Possible target-mediated adverse effects were identified via Open Targets Genetics^41,42^.

### Associations in DiscovEHR cohort

Individuals with a history of kidney stones were identified in the DiscovEHR cohort based on the presence of kidney stone ICD-10 codes (N20.0, N20.1, N20.2, N20.9 or N23) in their electronic health record. Unadjusted p values and odds ratios with 95% confidence intervals were calculated using the χ^2^ test and Cochran-Mantel-Haenszel statistic, respectively, to compare of the frequency of a kidney stone diagnosis between individuals carrying a variant of interest and those not carrying a variant of interest. Outpatient serum calcium and serum phosphorus lab values were collected and used to determine the median value. The average median value for serum calcium and serum phosphate in carriers and noncarriers was then compared. Log transformed serum phosphorus values were used for all statistical analysis as serum phosphorus values were not normally distributed. Effect sizes and p-values, adjusted for kidney stone diagnosis, were generated using logistic regression to assess how serum calcium and serum phosphorus levels are affected with the addition of 1 putative kidney stone-causing allele. This analysis was performed for each individual putative kidney stone-causing variant and with all three putative kidney stone-causing variants combined. Analysis was performed using SAS Enterprise Guide, version 8.3 (SAS Institute Inc, Cary, NC).

### Functional and structural characterization of DGKD variants

Individuals with kidney stone disease and a predicted ‘deleterious’ (SIFT) and ‘probably damaging’ (PolyPhen) *DGKD* missense variant with minor allele frequency <0.1% were identified in Genomics England 100,000 genomes (100KGP, Integrative Variant Analysis 2.0). Clinical data were obtained from Patient Explorer and referring clinicians.

Individuals with a history of kidney stone disease and those carrying *DGKD* rare variants were identified in the DiscovEHR cohort. Frequency of rare *DGKD* missense variant carriers with and without kidney stone disease were compared using χ^2^ test or Fisher exact test (as appropriate, Supplementary appendix). To determine relatedness between individuals within the DiscovEHR cohort, genome-wide identity by descent was used; once individuals were grouped into family networks, PRIMUS^43,44^ was applied (Supplementary appendix).

Conservation of DGKδ variants was assessed by aligning DGKδ orthologs with Clustal Omega. HEK293 cells were stably transfected to express CaSRs and FLAG-myc tagged *DGKD* or shRNA *DGKD*. Expression of DGKδ was confirmed via western blot analyses. The CaSR signals via pathways including Ras-Raf-MEK-ERK and intracellular calcium release; these signaling responses were assessed by serum response element (SRE) and nuclear factor of activated T cells-response element (NFAT-RE)-assays, respectively (Figure S8). SRE and NFAT-RE assays were undertaken following transfection of appropriate luciferase constructs and cellular responses to increasing extracellular calcium concentrations compared using 2-way ANOVA with Dunnett’s multiple-comparisons tests (GraphPad Prism v9). Cells were also treated with 5nM or 100nM cinacalcet.

### Three-dimensional modelling of DGK**δ** structure

The crystal structure of oligomeric DGKδ SAM domains has been determined (PDB 3BQ7^20^), and the structure of DGKδ isoform 2 predicted (AF-Q16760-F1-mod^45,46^, AlphaFold). The PyMOL Molecular Graphics System (Version 2.5.2, Schrödinger, LLC) was used for structural modelling based on these structures^47,48^. PyMOL Molecular Graphics System v2.5.2 and PyMod v3.0 were used to model the effects of DGKδ variants^47,49,50^.

## Data Availability

All data produced in the present study are available upon reasonable request to the authors

## Author contributions

Study conception: CE Lovegrove, M Goldsworthy, MV Holmes, D Furniss, RV Thakker, SA Howles

Data acquisition: CE Lovegrove, M Goldsworthy, J Haley, D Smelser, FM Hanan, A Mahajan, M Suri, O Sadeghi-Alavijeh, S Moochhala, D Gale, D Carey, SA Howles

Data analysis and interpretation: CE Lovegrove, M Goldsworthy, J Haley, D Smelser, C Gorvin, D Carey, MV Holmes, D Furniss, RV Thakker, SA Howles

Manuscript preparation and review: CE Lovegrove, M Goldsworthy, J Haley, D Smelser, C Gorvin, FM Hanan M Suri, O Sadeghi-Alavijeh, S Moochhala, D Gale, D Carey, MV Holmes, D Furniss, RV Thakker, SA Howles

Final approval of manuscript: CE Lovegrove, M Goldsworthy, J Haley, D Smelser, C Gorvin, FM Hanan, A Mahajan, M Suri, O Sadeghi-Alavijeh, S Moochhala, D Gale, D Carey, MV Holmes, D Furniss, RV Thakker, SA Howles

### Acknowledgements

We acknowledge the contribution of participants and investigators of the UK Biobank. This research was made possible through access to the data and findings generated by the 100,000 Genomes Project (100KGP), which is managed by Genomics England Ltd (a wholly owned company of the Department of Health and Social Care). The 100KGP is funded by the National Institute for Health Research and the National Health Service of England. The Wellcome Trust, Cancer Research UK, and the Medical Research Council (MRC) have also funded research infrastructure. The 100KGP uses data provided by patients and collected by the National Health Service as part of their care and support. The authors gratefully acknowledge the participation of the patients and their families recruited to the 100KGP. We would also like to thank Mark McCarthy for allowing us access to the calcium and phosphate GWAS results that underlie much of the work in this manuscript.

## Rights retention statement

*For the purpose of Open Access, the author has applied a CC BY public copyright licence to any Author Accepted Manuscript (AAM) version arising from this submission.*

