## Supplementary appendix for "Genetic variants predisposing to increased risk of kidney stone disease"

This appendix has been provided by the authors to give readers additional information about the work.

Supplementary Methods

**Genome-wide association study of kidney stone disease in UK Biobank**

Genotype phasing and imputation in the UK Biobank study using UK-BiLEVE and UK-Biobank Axiom Arrays has been previously described^1^. This included 92,693,895 autosomal SNPs, short indels and large structural variants. R version 4.2.0 and PLINKv2.0 were used for quality control. Single nucleotide polymorphisms with a call rate < 90% were removed, accounting for the two different genotyping platforms used to genotype individuals. SNP-level QC excluded SNPs with Hardy–Weinberg equilibrium p<10^−4^, <98% call rate, and minor allele frequency (MAF) <1%. Following QC, data from 547,011 autosomal genotyped and 8,397,548 imputed variants were considered.

An hg19 reference genetic map and a reference linkage disequilibrium score file for European ancestry were used. Quantile–quantile and Manhattan plots were generated in FUMA^2^.

Heritability estimates were calculated using LD score regression (LDSC) v1.0.1^3,4^. Analyses were restricted to variants in HapMap3^5^ and used LD Scores computed using 1000 Genomes European data^6^. A population prevalence approximation of 10% was used in liability transformation. We used GCTA (Genome-Wide Complex Trait Analysis) software Version 1.94.1 to perform step-wise approximate conditional and joint analysis with the same UK Biobank LD reference panel as was used in the UK Biobank KSD GWAS^7,8^. Where there was a single signal of association at a locus (a chromosomal region with adjacent pairs of KSD-associated SNPs <1Mb apart^7,9^), we defined the index SNP as the lead SNP from unconditional analysis. For loci with multiple association signals, we defined the index SNP as that with the lowest P value in conditional approximate analysis.

**Genome-wide association study of albumin-adjusted serum calcium and serum phosphate concentrations in UK Biobank**

Serum albumin-adjusted calcium concentrations for UK Biobank participants were derived using the following equation: adjusted calcium (mmol/L)=total Calcium(mmol/L) + 0.0177 *(46.3 – albumin (g/L)). Data from participants with eGFR(MDRD) <60ml/min/1.73m2 and 25-OH vitamin D concentrations <30nmol/L were excluded from association analyses for both serum phosphate concentrations and serum albumin-adjusted calcium concentrations. Analyses were performed using genotyped and imputed variants from the UK Biobank. Genotyping was undertaken using UK-BiLEVE and UK-Biobank Axiom Arrays. Phenotypes were inverse normalized with additional adjustments for array, age, and sex. Analyses were undertaken in individuals of European ancestry using BOLT-LMM to account for population sub-structure and cryptic relatedness. Imputed SNPs of minor allele frequency (MAF) <1% and of imputation quality score <0.3 were excluded from analyses. Lead SNPs were identified from unconditional analyses and loci defined as ±500kb surrounding each SNP. Overlapping loci were merged as one locus. GCTA was used to perform a stepwise model selection procedure to select independently-associated SNPs within each 1Mb region of significance p<5x10-9. Directly genotyped variants underwent stringent quality control checks, including call rate per array, manual cluster plot checks and status in Gnomad. Only variants with MAF <1% and the coding or loss-of-function annotations of “missense variant”, “stop gain”, “frameshift variant”, “splice acceptor variant”, “splice donor variant”, “splice region variant”, “start lost”, or “stop lost” were included. A significance threshold of p<5x10-6 was used to identify directly genotyped SNPs with significant associations with each phenotype.

**Mendelian Randomization**

We undertook Mendelian randomization (MR), using the TwoSample MR package in R, to estimate the effects of genetically-instrumented mineral metabolism traits on odds of kidney stone disease at loci significantly associated with serum adjusted-calcium or phosphate from GWAS in the UK Biobank (Supplementary data 1)^10,11^. MR assumes that instrumental variables (IVs) are associated with the exposure variable (relevance); there are no unmeasured confounding relationships (exchangeability); and variants are associated with the outcome only through changes in the exposure variable (exclusion restriction)^12^. We extracted significant genetic variants ±500kbp of lead, independent SNPs associated with adjusted calcium or phosphate at GWAS (p<5x10^-8^) and performed clumping according to *r^2^*<0.1 using a European population reference. Exposure IVs were harmonized with outcome IVs from GWAS of kidney stone disease in the UK Biobank. Allele frequencies were used to infer positive strand alleles for palindromic IVs. Where harmonization was not possible and the positive strand alleles remained ambiguous, IVs were omitted. We performed a power calculation using https://sb452.shinyapps.io/overlap/ to calculate the minimum and maximum effects that we had 80% statistical power to detect (Table S3)^13,14^.

To assess the plausibility of the core IV assumptions, variance in exposure trait explained by the genetic variants (*R^2^*) was calculated: *R^2^* = [2 × MAF × (1 – MAF) × β^2^], where MAF is the minor allele frequency and β is the log-odds of the SNP^15^. Mean F statistics for exposure IVs were calculated using the following formulae where the genetic association with the risk factor ($â$) is in standard deviation units, *MAF* is the minor allele frequency, *N* is the sample size for the IV–outcome association, and *K* is the number of IVs^16^:

$$R^{2}=2â^{2}x MAF x (1-MAF)$$

$$\text{F=}\frac{R^{2}(N-1-K)}{\left( 1-R^{2} \right)K}$$

**Identifying potential target-mediated adverse effects of gene targets**

Possible off-target effects of putative therapeutics were identified by collating phenotypes associated with variants linked to these genes (p<5x10-8) via the Open Targets Genetics portal (https://genetics.opentargets.org/, accessed 30/01/2024)^17,18^.

**Genotyping in the DiscovEHR cohort**

Genotype was determined using DNA extracted from MyCode participant blood or saliva samples collected as part of the DiscovEHR collaboration^19^. Regeneron Genetic Center (RGC, Tarrytown, NY) performed genotyping using the Illumina Infinium Global Screening Array GSA-24v2-0_A2 (Illumina, Inc, San Diego, CA) then filtered for minor allele frequency (MAF) >1%, Hardy-Weinberg Equilibrium (HWE) p-value >1x10^-15^ and site missingness <1%. Data was uploaded by batch to the TOPMed Imputation Server (<https://imputation.biodatacatalyst.nhlbi.nih.gov>) for genotype imputation using MINIMAC4 and the TOPMed reference panel.

**Rare variant analyses**

Individuals with a history of kidney stones were identified in the DiscovEHR cohort based on the presence of an N20.0, N20.1, N20.2, N20.9 or N23 ICD10 in their electronic health record. DGKD rare variant carriers were identified using data from exome sequencing of DNA extracted from the blood of each DiscovEHR participant. Exome data was filtered using the following quality control metrics: MAF<0.1, combined depth of ≥10 for indels, quality by depth >3 and combined depth of ≥7 for single nucleotide variants, alternate allelic balance >15% (single nucleotide variants) or >20% (indels), and ≥5 alternate reads. Variants were annotated with variant type (e.g. missense), human genome variation, and variant definition (including position, gene name, predicted protein coding alterations). Frequency of rare DGKD missense variant carriers with and without a kidney stone diagnosis was compared using χ^2^ test or Fisher exact test (as appropriate). Unadjusted and adjusted odds ratios and 95% confidence intervals were calculated using the Cochran-Mantel-Haenszel statistics or logistic regression to test for significant associations between these variables and a kidney stone diagnosis. Analysis was performed using SAS Enterprise Guide, version 8.3 (SAS Institute Inc, Cary, NC).

**Relationship and pedigrees in DiscovEHR cohort**

To determine the relatedness between individuals within the DiscovEHR cohort, genome-wide identity by descent (IBD) was used^20^. Ancestral class (Admixed American, African, East Asian, European, South Asian, and unknown) was determined using principal components and the HapMap3 dataset. High quality common variants (MAF >0.1, missingness 0.05, and expected heterozygosity rates) and high quality samples (percent 20x coverage <0.75) were used to calculate pairwise IBD within ancestral groups. A PI_HAT threshold 0.1875 for 2^nd^ degree relationships and 0.3 for 1^st^ degree relationships. Individuals were then grouped into family networks and run through PRIMUS^21^ for improved IBD estimates to determine the relationships within each family network.

**Functional analyses**

Functional studies were undertaken using HEK293 cells (ATCC^®^ CRL-1573™) transfected using the FLP-In system to express calcium-sensing receptors (CaSRs) (HEK-FLP-In CaSR cells). For overexpression studies a Myc-DDK tagged DGKDv2 cDNA (NM_152879) in the pCMV6_Entry vector (Cat No. RC217053) clone was purchased from Origene. Point mutations were introduced into this clone using QuickChange Lightning Site Directed Mutagenesis kit (Agilent) according to the manufacturer’s instructions to produce constructs containing the Ile91Val, His190Gln, Ile221Asn, Thr319Ala, Val464Ile, Arg900His, and Arg 1181Trp variants. Constructs were sequenced to confirm presence of variants prior to transfection into HEK FLP-In CaSR cells and overexpression (OE) stable cell lines selected for by growth in Geneticin media. Cell lines were subsequently maintained in DMEM-Glutamax media (Thermo Fisher Scientific) with 10% foetal bovine serum (FBS) (Gibco) and 400 μg/ml geneticin (Thermo Fisher Scientific) and 200 μg/ml hygromycin (Invitrogen) at 37 °C, 5% CO_2_. These cell lines were utilized in subsequent serum-response element (SRE) and nuclear factor of activated cells (NFAT) assays.

Expression of DGKδ and CaSR was confirmed via western blot analyses. Western blot analyses were undertaken using anti-cMyc (A190-105P; ThermoFisher Scientific; 1: 3000), anti-DGKD (GTX87254; GeneTex; 1:1000), anti-CaSR (5C10, ADD; ab19347; Abcam; 1: 6000), and anti−α−Tubulin (T5168; Sigma; 1: 3000) antibodies. The western blots were visualized using an Immuno-Star Western C kit (Bio-Rad) on a Bio-Rad Chemidoc XRS + system.

DGKD Human shRNA Plasmid kit (Locus ID8527) (Cat No. TF313492) containing 4 differing shRNAs in pRFP-C-RS Vector was utilized to generate stable knockdown (KD) cell lines in HEK-FLP-In CaSR cells according to the manufacturer’s instructions. Stable cell lines were maintained in DMEM-Glutamax media (Thermo Fisher Scientific) with 10% FBS (Gibco) and 1 μg/ml Puromycin (Thermo Fisher Scientific) and 200 μg/ml hygromycin (Invitrogen) at 37 °C, 5% CO_2_. These cell lines were utilized in subsequent SRE and NFAT assays.

Successful KD of DGKD and maintenance of CaSR expression was confirmed via quantitative reverse transcriptase PCR (qRT-PCR) and western blot analyses. qRT-PCR analyses were performed in quadruplicate using Power SYBR Green Cells-to-CT™ Kit (Life Technologies), *DGKD, CASR, PGK1*, *GAPDH*, *TUB1A*, *CDNK1B* specific primers (Qiagen), and a Rotor-Gene Q real-time cycler (Qiagen Inc, Valencia, CA). Samples were normalized to a geometric mean of four housekeeper genes: *PGK1*, *GAPDH*, *TUB1A*, *CDNK1B*.

To perform SRE response assays, HEK-FLP-In CaSR- DGKδ OE or KD cells were plated into 96 well plates and transfected with an SRE reporter assay plasmid (Promega) using lipofectamine 2000 according to the manufacturer’s instructions. Thirty-six hours post transfection cells were incubated in 0.05% FBS media with 0.45 mM calcium for 12 hours, reducing extracellular calcium concentration and thus inducing basal cellular CaSR-mediated responses whilst maintaining cellular viability. At 48 hours post transfection the media was changed to varying concentrations of extracellular calcium (0.1–5 mM), with either 5 nM cinacalcet, 100nM cinacalcet, or equivalent volume of dimethylsulfoxide (DMSO) (final concentration of DMSO 0.0001%), and the cells were incubated for a further 4 hours at 37 °C. Cinacalcet (AMG-073 HCL) was obtained from Cambridge Bioscience (catalog CAY16042) and dissolved in DMSO prior to use in in vitro studies. Cells were lysed and luciferase activity measured using Luciferase Assay System (Promega) on a PHERAstar microplate reader (BMG Labtech). Assays were performed in > 4 biological replicates (independently transfected wells, performed on at least 4 different days). Nonlinear regression of concentration-response curves was performed with GraphPad Prismv9 for determinations of maximal response.

To perform NFAT response assays, HEK-FLP-In CaSR- DGKδ OE or KD cells were plated into 96 well plates and transfected with an NFAT reporter assay plasmid (Promega) using lipofectamine 2000 according to the manufacturer’s instructions. 36 hours post transfection cells were incubated in 0.05% FBS media with 0.45 mM calcium for 12 hours, reducing extracellular calcium concentration and thus inducing basal cellular CaSR-mediated responses whilst maintaining cellular viability. At 48 hours post transfection the media was changed to varying concentrations of extracellular calcium (0.1–10 mM) and the cells were incubated for a further 4 hours at 37 °C. Cells were lysed and luciferase activity measured using Luciferase Assay System (Promega) on a PHERAstar microplate reader (BMG Labtech). Assays were performed in > 4 biological replicates (independently transfected wells, performed on at least 4 different days). Nonlinear regression of concentration-response curves was performed with GraphPad Prismv9 for determinations of maximal response.

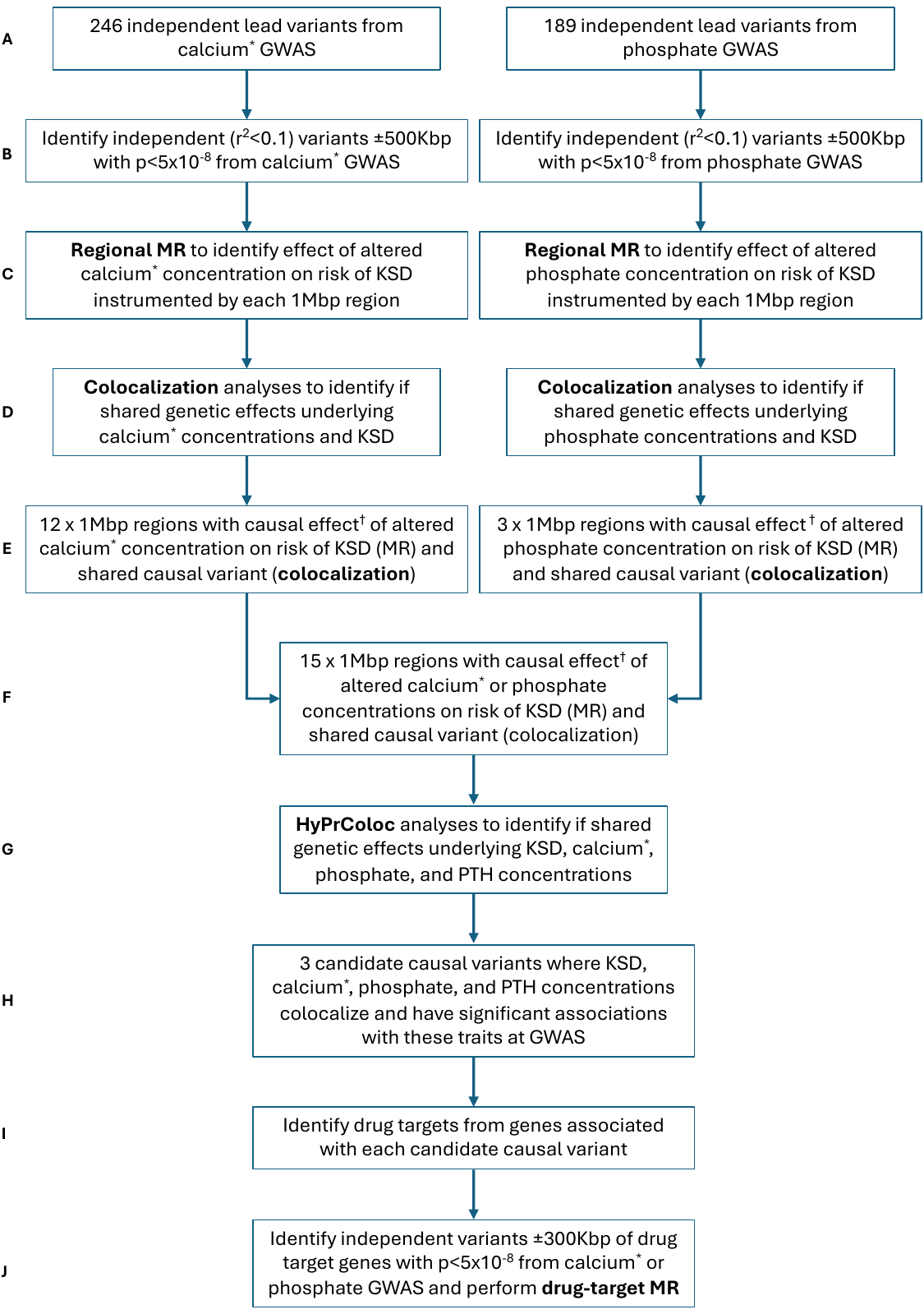

Supplementary Figure S1: Study design to identify genetic variants predisposing to increased risk of kidney stone disease

**A, B:** Independent (r2<0.1) genetic variants ±500kbp of lead independent variants from serum albumin-adjusted calcium or phosphate genome-wide association studies (GWAS) significantly (p<5x10-8) associated with serum albumin-adjusted calcium or phosphate concentrations were selected for use as instrumental variables (IVs). **C**: Mendelian randomization (MR) was performed using each of the identified IVs to instrument the effects of alterations in the biochemical exposure on risk of kidney stone disease (KSD) using UK Biobank GWAS as the outcome. **D**: Colocalization analyses were performed. **E**: Regions with significant MR results (after p-value adjustment using the false-discovery rate method) and evidence of colocalization were identified. **F,G,H**: HyPrColoc was undertaken to assess if there was colocalization between kidney stone disease and serum albumin-adjusted serum calcium, phosphate, and parathyroid hormone (PTH) concentrations and identity candidate causal variants. **I:** Drug-targets from the genes associated with candidate causal variants were identified. **J**: Drug-target MR was performed to assess the potential utility of modulating drug targets to prevent KSD, selecting genetic variants for use as IVs within 300kbp of genes of interest. GWAS= genome-wide association study; Kbp=kilo-base pairs; KSD= kidney stone disease; Mbp= mega-base pairs; MR= Mendelian randomization; PTH= parathyroid hormone; ^*^albumin-adjusted serum calcium concentration; ^†^instrumental variable comprising 3 or more genetic variants.

**A**
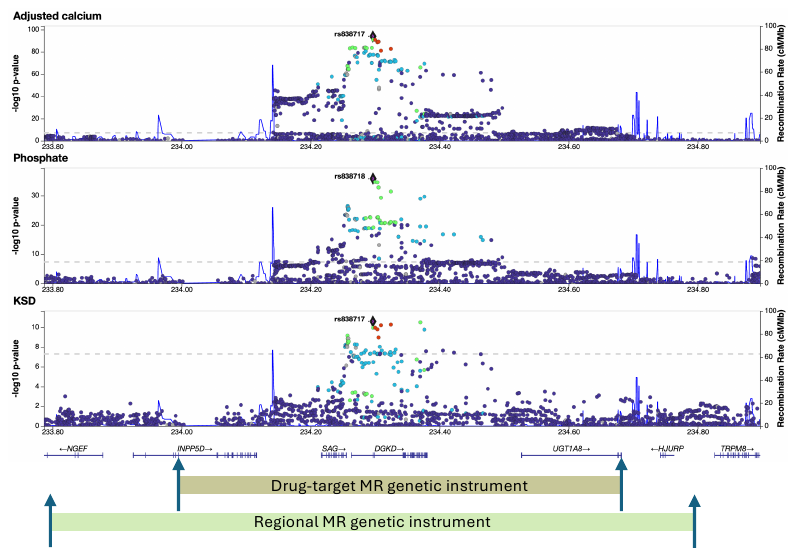

**B**
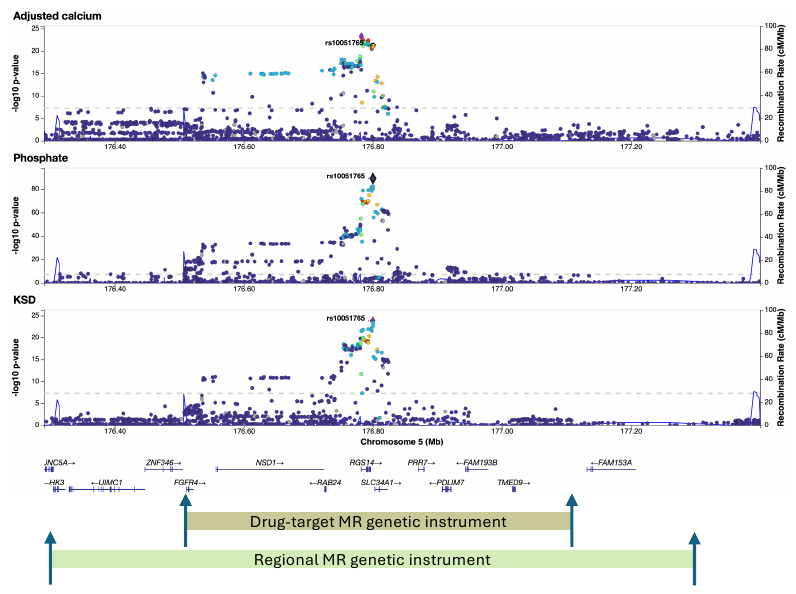

**C**
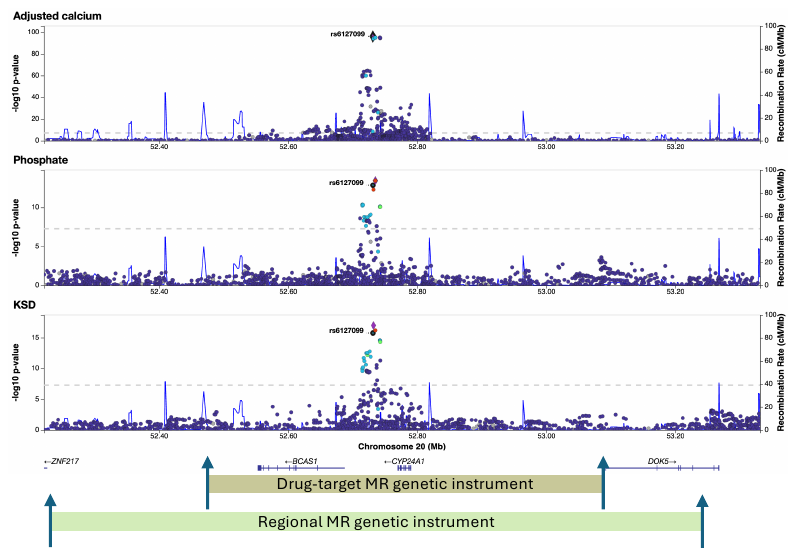

Supplementary Figure S2: Locus zoom plots showing genetic coordinates from which regional and drug-target Mendelian randomization instrumental variables were selected

Green boxes denote genomic regions from which genetic variants were selected for regional Mendelian randomization (MR); these variants were within a 500kbp window either side of lead independent variants from serum albumin-adjusted calcium or phosphate genome-wide association studies. Brown boxes denote genomic regions from which genetic variants were selected for drug-target MR; these variants were within a 300kbp window either side of the gene coordinates for genes associated with candidate causal variants identified from MR and colocalization analyses. Candidate causal variants are highlighted in each locus zoom plot in the **A**: *DGKD*, **B**: *SLC34A1*, and **C**: *CYP24A1* regions using data from genome-wide association studies of albumin-adjusted serum calcium, phosphate, and kidney stone disease (KSD).

A
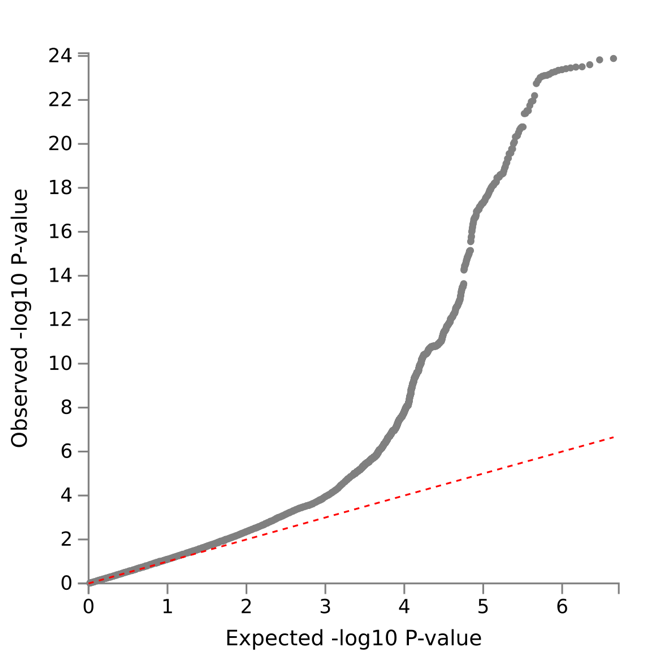

Supplementary Figure S3: Quantile-quantile plot of observed vs. expected p-values for genome-wide association study (GWAS) of 11,186 kidney stone disease cases and 390,488 controls in the UK Biobank.

The λGC demonstrated some inflation (1.15), but the LD score regression (LDSC) intercept of 1.01, with an attenuation ratio of 0.09 indicated that the inflation was largely due to polygenicity and the large sample size.

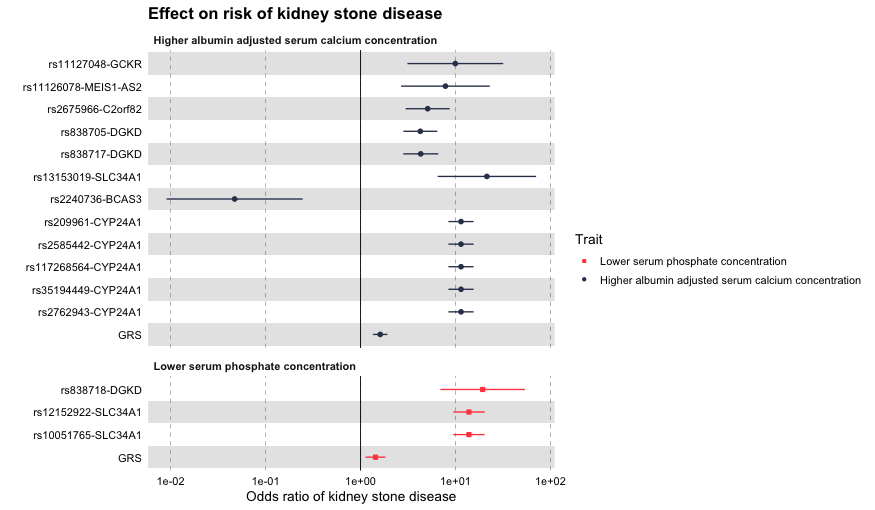

Supplementary Figure S4: Forest plot demonstrating Mendelian randomization estimates for predicted causal variants (±500kb) on liability to kidney stone disease in the UK Biobank.

GRS=overall Mendelian randomization estimate for trait on risk of kidney stone disease

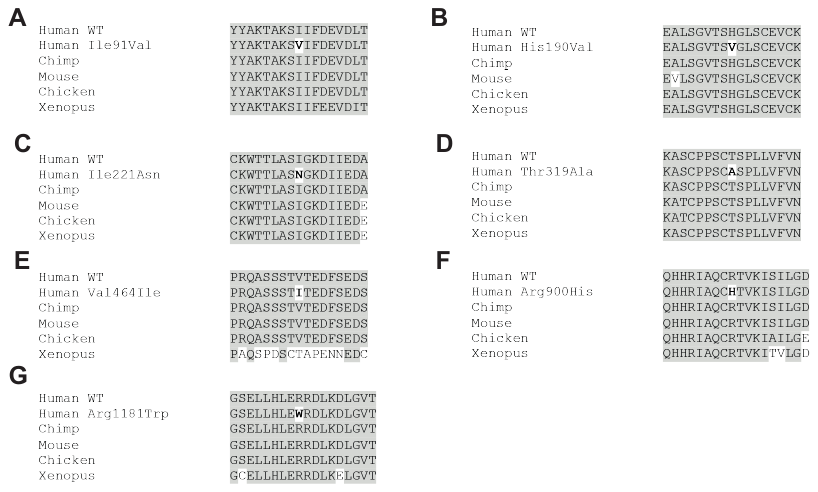

Supplementary Figure S5: Multiple protein sequence alignment of DGKδ

Evolutionary conservation of residues implicated in kidney stone disease in orthologs. Conserved residues are shaded grey.

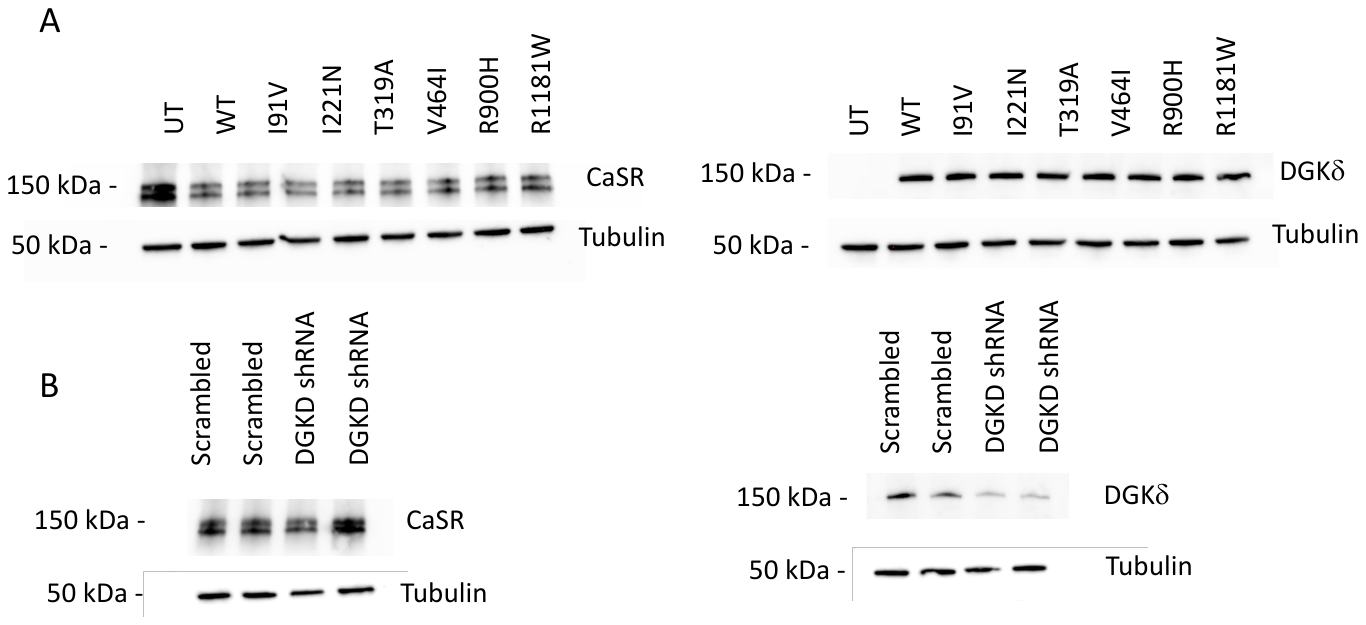

Supplementary Figure S6: Expression of DGKδ variants in HEK293 cells.

**A :** Representative western blot of lysates from HEK-CaSR stably transfected with Myc-tagged DGKD; α−Tubulin was used as a loading control. Anti-CaSR, anti-tubulin, and anti-Myc antibodies were used. UT-untransfected cells, WT- wildtype. **B:** Representative western blot of lysates from HEK-CaSR cells treated with scrambled or DGKD shRNA; α−Tubulin was used as a loading control. Anti-CaSR, anti-tubulin, and anti-DGKD antibodies were used.

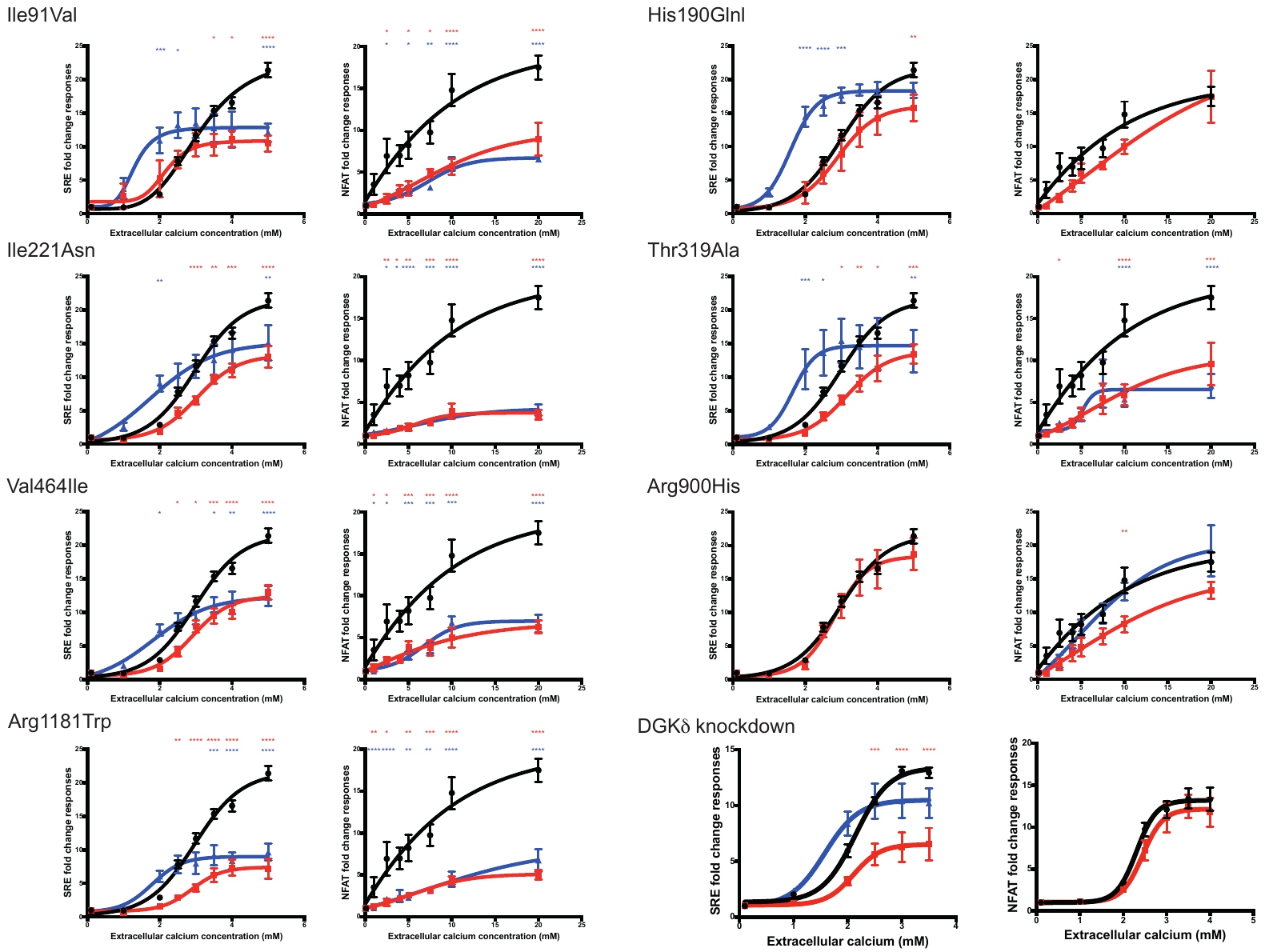

Supplementary Figure S7: Functional characterization of kidney stone-associated DGKδ variants. CaSR-mediated SRE and NFAT responses to changes in extracellular calcium concentration [Ca2+]e and effect of 100nM cinacalcet treatment (missense variants) and 5nM cinacalcet (DGKδ knockdown) in HEK-CaSR-DGKδ cells transfected with wild-type or kidney stone-associated variants Ile91Val, His190Gln, Ile221Asn, Thr319Ala, Val464Ile, Arg900His, and Arg 1181Trp, and in HEK-CaSR cells following DGKδ knockdown. The responses ± standard error of the mean (SEM) are shown for n>4 biologically independent experiments. Transfection with kidney stone-associated DGKD variants and DGKδ knockdown led to a reduction in responses (red line) compared to cells transfected with wild-type DGKD (black line). Treatment with cinacalcet increased SRE-mediated responses but had no effect on NFAT responses (blue line). Two-way ANOVA with Dunnet’s correction for multiple comparisons was used to compare points on dose response curve with reference to wild-type. Data are shown as mean ± SEM with *p<0.05, **p < 0.01, ***p<0.001, ****p < 0.0001.

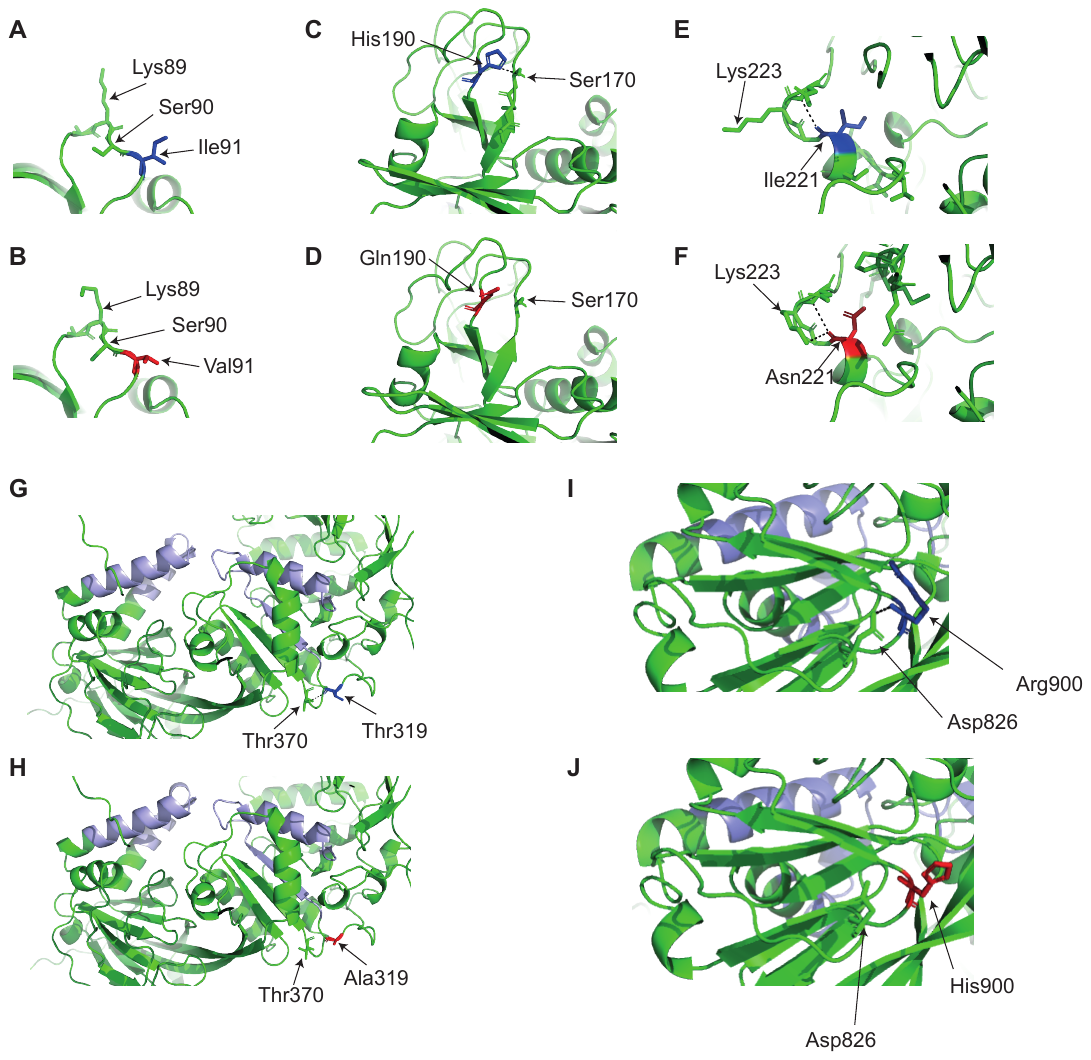

Supplementary Figure S8: Predicted effects of kidney stone-associated DGKδ variants based on the predicted structure of DGKδ isoform 2, (AF-Q16760-F1-mod^22,23^ AlphaFold)

**A and B:** Residue Ile91 (wild-type, dark blue) is within the β3-β4 linker of the DGKδ predicted pleckstrin homology (PH) domain; the mutation Val91 (red); is predicted to result in an altered confirmation of residues Lys89 and Ser90. Residues in an equivalent position have been shown to be important for binding of the PH domain of phospholipase C beta to inositol trisphosphate (Ferguson et al. Cell 83:1037-1046. 1995. PMID: 8521504.) **C and D:** His190 wild-type, (dark blue) and Gln190 (red); the mutation Gln190 is predicted to result in the loss of a polar contact (dashed black line, Ser170) between two β sheets. Studies of the C1 domain of Protein Kinase Cδ suggest that these two β sheets are pulled apart to enable ligand binding. **E and F:** Ile221 (wild-type, dark blue) and Asn221 (red); the mutation Asn221 is predicted to result in an additional polar contact (dashed black line) with Lys223 and alteration in residue orientation on comparison to the Ile221 protein. These residues are within the C1 domain. **G and H:** Thr319 (wild-type, dark blue) and Ala319 (red); the mutation Ala319 is predicted to result in the loss of a polar contact (dashed black line) with Thr370. Ala319 is in close proximity to the ATP-binding pocket (light blue), this change in polar contacts may alter the conformation of this cleft. **I and J:** Arg900 (wild-type, dark blue) and His900 (red); the mutation His900 is predicted to result in the loss of a polar contact (dashed black line) with Asp826. Asp826 forms is linked to the accessory domain ATP-binding motif (light blue).

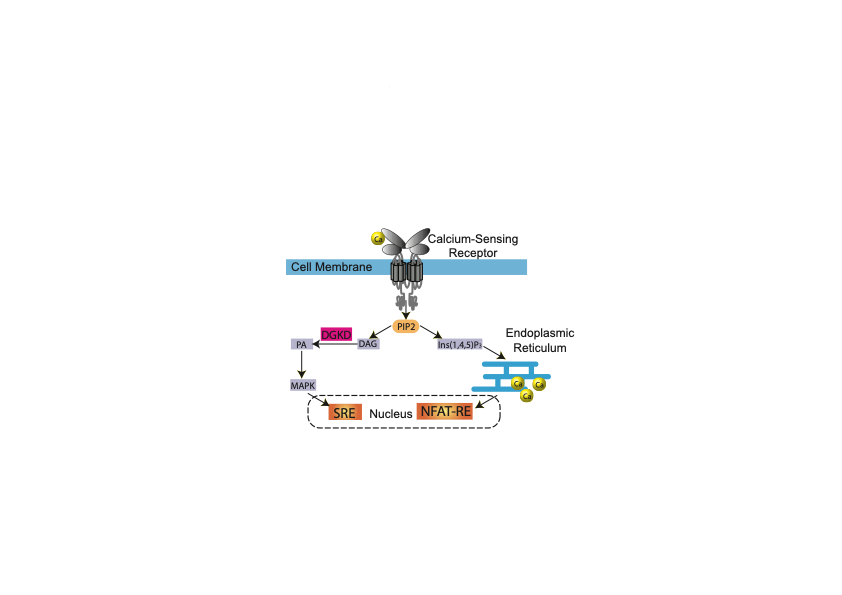

Supplementary Figure S9. Calcium sensing receptor signaling pathway**.** Binding of calcium (yellow) to the extracellular bilobed venus fly-trap domain of the CaSR (light grey) results in Gα11 dependent stimulation of phospholipase C-β, which catalyzes the formation of inositol 1,4,5-trisphosphate (Ins(1,4,5)P_3_) and diacylglycerol (DAG) from phosphatidylinositol 4,5-bisphosphate (PIP2). An accumulation of Ins(1,4,5)P_3_ mediates calcium mobilization into the cytosol from intracellular stores, whereas DAG activates the phospho-extracellular signal regulated kinase (pERK) arm of the mitogen activated protein kinase (MAPK) cascade via the production of phosphatidic acid (PA). Gene transcription mediated by intracellular calcium and MAPK signaling can be measured using nuclear factor of activated T-cells response element (NFAT-RE) and serum response element (SRE) containing luciferase reporter constructs, respectively.

Supplementary Table S1: Variants associated with kidney stone disease at genome-wide association study in the UK Biobank

| Variant | CHR | POS | Candidate gene | EA | NEA | EAF | Conditional OR (95% CI) | Conditional P |
| --- | --- | --- | --- | --- | --- | --- | --- | --- |
| rs77362499 | 1 | 21836204 | *ALPL* | G | C | 0.11 | 1.23 (1.17-1.28) | 2.70x10-20 |
| rs1256332 | 1 | 21893344 | *ALPL* | A | C | 0.16 | 1.17 (1.13-1.21) | 3.81x10-17 |
| rs838717 | 2 | 234296444 | *DGKD* | G | A | 0.43 | 1.1 (1.07-1.13) | 2.60x10-11 |
| rs10051765 | 5 | 176799992 | *SLC34A1* | C | T | 0.33 | 1.16 (1.13-1.19) | 1.30x10-24 |
| rs7774646 | 6 | 30760454 | *FLOT1** | T | G | 0.17 | 1.11 (1.07-1.15) | 2.30x10-9 |
| rs1155347 | 6 | 39146230 | *KCNK5* | C | T | 0.22 | 1.13 (1.09-1.16) | 4.40x10-13 |
| rs74348938 | 6 | 160614649 | *SLC22A2* | A | G | 0.03 | 1.28 (1.18-1.38) | 5.30x10-10 |
| rs5883088 | 7 | 27610433 | *HIBADH* | G | GC | 0.33 | 1.11 (1.08-1.14) | 2.80x10-12 |
| rs4252512 | 7 | 142605221 | *TRPV5* | C | T | 0.02 | 1.34 (1.22-1.49) | 7.70x10-9 |
| rs7124537 | 11 | 10505718 | *AMPD3* | C | A | 0.09 | 1.15 (1.1-1.2) | 5.40x10-9 |
| rs1182959 | 13 | 42674601 | *DGKH* | A | G | 0.18 | 1.14 (1.1-1.18) | 1.30x10-13 |
| rs76594840 | 15 | 47900177 | *SEMA6D** | T | C | 0.92 | 1.15 (1.09-1.21) | 3.80x10-8 |
| rs112414002 | 16 | 16284360 | *ABCC6** | C | T | 0.97 | 1.27 (1.16-1.38) | 4.40x10-8 |
| rs77924615 | 16 | 20392332 | *UMOD* | A | G | 0.20 | 1.15 (1.11-1.19) | 7.50x10-16 |
| rs9902482 | 17 | 70356993 | *SOX9* | C | T | 0.50 | 1.08 (1.05-1.11) | 1.10x10-8 |
| rs17216707 | 20 | 52732362 | *CYP24A1* | T | C | 0.81 | 1.16 (1.12-1.2) | 9.70x10-18 |
| rs219772 | 21 | 37835347 | *CLDN14* | A | T | 0.74 | 1.17 (1.13-1.21) | 2.50x10-24 |
| rs13054904 | 22 | 23410918 | *GNAZ* | A | T | 0.26 | 1.14 (1.11-1.18) | 7.00x10-18 |

CHR = chromosome; POS = position based on NCBI Genome Build 37 (hg19); EA= effect allele; NEA= Non-effect allele; EAF= effect allele frequency; OR= odds ratio; P= P-value; 95% CI= 95% confidence interval

Supplementary Table S2: Heritability estimates for GWAS in UK Biobank

|  | UK Biobank GWAS |
| --- | --- |
| N SNPs | 1,154,335 |
| N cases | 11,186 |
| N controls | 390,488 |
| *h*2 _SNP_- liability scale  (SE) | 0.19  (0.02) |
| LDSC intercept  (SE) | 1.01  (0.01) |
| LDSC ratio  (SE) | 0.09  (0.5) |
| Mean χ^2^ | 1.16 |

Estimates are calculated using LD-score regression (LDSC) at a population prevalence of 10%. *h*2 _SNP_= mean SNP-based heritability; N SNPs= number of SNPs analyzed; N cases= number of cases; N controls= number of controls; SE= standard error

Supplementary Table S3: Estimate of bias for Mendelian randomization with sample overlap in the UK Biobank

| Mineral metabolism trait | GWAS sample size | Maximum number of instruments in MR analyses | Estimate of bias |
| --- | --- | --- | --- |
| Adjusted calcium | 308,679 | 24 | 0.011 |
| Phosphate | 308,110 | 20 | 0.001 |

The maximum number of instruments was used to derive the most pessimistic estimate of bias. Results are shown for a Type 1 error rate of 0.05.

Supplementary Table S4: Causal effects of lead variants from mineral metabolism GWAS (±500kb) with evidence of colocalization on kidney stone risk

| Trait | Lead SNP from mineral metabolism genome wide association study in UK Biobank | | | | Mendelian randomization with kidney stone disease | | | | | | | | | Colocalization analyses with kidney stone disease | | | |
| --- | --- | --- | --- | --- | --- | --- | --- | --- | --- | --- | --- | --- | --- | --- | --- | --- | --- |
|  |  |  |  |  | Instrumental variables | | Estimate | | Intercept | | Heterogeneity | | |  |  |  |  |
|  | SNP | Gene | CHR | POS | N SNPs | Mean R^2^ (SD) | OR (95% CI) | FDR-P | Beta (SE) | P | Q | Q_df | P | N SNPs | PP H4 | Candidate causal SNP | SNP PP |
| Increasing adjusted calcium | rs11127048 | *GCKR* | 2 | 27752463 | 3 | 1.25x10-4 (9.08x10-5) | 9.94 (3.11-31.76) | 1.12x10^-3^ | -0.04 (0.03) | 0.47 | 2.09 | 2 | 0.35 | 1750 | 0.87 | rs1260326 | 0.67 |
| Increasing adjusted calcium | rs11126078 | *MEIS1* | 2 | 66656402 | 6 | 6.80x10-5 (2.08x10-5) | 7.83 (2.67-22.95) | 1.63x10^-3^ | 0.01 (0.04) | 0.83 | 0.41 | 5 | 1.00 | 3140 | 0.83 | rs2049019 | 0.21 |
| Increasing adjusted calcium | rs2675966 | *C2orf82* | 2 | 233735543 | 8 | 2.03x10-4 (2.56x10-4) | 5.09 (2.98-8.68) | 7.42x10^-8^ | -0.01 (0.03) | 0.75 | 6.16 | 7 | 0.52 | 3600 | 0.96 | rs11891546 | 1.00 |
| Increasing adjusted calcium | rs838705 | *DGKD* | 2 | 234273242 | 19 | 1.99x10-4 (2.88x10-4) | 4.26 (2.82-6.43) | 2.17x10-^10^ | -0.02 (0.02) | 0.24 | 24.96 | 18 | 0.13 | 3400 | 1.00 | rs838717 | 1.00 |
| Increasing adjusted calcium | rs838717 | *DGKD* | 2 | 234296444 | 18 | 2.07x10-4 (2.94x10-4) | 4.3 (2.81-6.58) | 5.61x10^-10^ | -0.02 (0.02) | 0.29 | 24.69 | 17 | 0.10 | 3360 | 1.00 | rs838717 | 1.00 |
| Increasing adjusted calcium | rs13153019 | *SLC34A1* | 5 | 176782218 | 4 | 1.56x10-4 (1.12x10-4) | 21.38 (6.48-70.52) | 1.19x10^-5^ | -0.02 (0.08) | 0.8 | 6.28 | 3 | 0.10 | 2390 | 0.89 | rs10051765 | 0.55 |
| Increasing adjusted calcium | rs2240736 | *BCAS3* | 17 | 59485393 | 6 | 1.14x10-4 (7.37x10-5) | 0.05 (0.01-0.25) | 2.37x10^-3^ | 0.01 (0.09) | 0.92 | 18.60 | 5 | 0.00 | 1810 | 0.97 | rs9905274 | 0.48 |
| Increasing adjusted calcium | rs209961 | *CYP24A1* | 20 | 52715154 | 24 | 2.54x10-4 (2.84x10-4) | 11.42 (8.41-15.5) | 2.45x10^-53^ | -0.02 (0.01) | 0.09 | 29.93 | 23 | 0.15 | 3630 | 0.98 | rs6127099 | 0.98 |
| Increasing adjusted calcium | rs2585442 | *CYP24A1* | 20 | 52737123 | 24 | 2.54x10-4 (2.84x10-4) | 11.42 (8.41-15.5) | 2.45x10^-53^ | -0.02 (0.01) | 0.09 | 29.93 | 23 | 0.15 | 3670 | 0.98 | rs6127099 | 0.98 |
| Increasing adjusted calcium | rs117268564 | *CYP24A1* | 20 | 52738434 | 24 | 2.54x10-4 (2.84x10-4) | 11.42 (8.41-15.5) | 2.45x10^-53^ | -0.02 (0.01) | 0.09 | 29.93 | 23 | 0.15 | 3670 | 0.98 | rs6127099 | 0.98 |
| Increasing adjusted calcium | rs35194449 | *CYP24A1* | 20 | 52742047 | 24 | 2.54x10-4 (2.84x10-4) | 11.42 (8.41-15.5) | 2.45x10^-53^ | -0.02 (0.01) | 0.09 | 29.93 | 23 | 0.15 | 3670 | 0.98 | rs6127099 | 0.98 |
| Increasing adjusted calcium | rs2762943 | *CYP24A1* | 20 | 52790786 | 24 | 2.54x10-4 (2.84x10-4) | 11.42 (8.41-15.5) | 2.45x10^-53^ | -0.02 (0.01) | 0.09 | 29.93 | 23 | 0.15 | 3750 | 0.98 | rs6127099 | 0.98 |
| Decreasing phosphate | rs838718 | *DGKD* | 2 | 234296650 | 4 | 1.96x10-4 (1.87x10-4) | 19.28  (6.92-53.68) | 8.91x10^-7^ | 0.00 (0.05) | 0.98 | 6.99 | 3 | 0.07 | 3370 | 0.99 | rs838718 | 0.48 |
| Decreasing phosphate | rs12152922 | *SLC34A1* | 5 | 176645599 | 9 | 2.73x10-4 (3.56x10-4) | 13.83 (9.44-20.27) | 2.42x10^-39^ | 0.03 (0.01) | 0.11 | 7.54 | 8 | 0.48 | 2580 | 1.00 | rs10051765 | 0.82 |
| Decreasing phosphate | rs10051765 | *SLC34A1* | 5 | 176799992 | 9 | 2.73x10-4 (3.56x10-4) | 13.83 (9.44-20.27) | 2.42x10^-39^ | 0.03 (0.01) | 0.11 | 7.54 | 8 | 0.48 | 2360 | 1.00 | rs10051765 | 0.82 |

CHR= chromosome; POS= position based on NCBI Genome Build 37 (hg19); N SNPs= number of SNPs; FDR-P= P-value adjusted for 5% false discovery rate; OR= odds ratio for kidney stone disease per genetically-instrumented standard deviation increase in trait; P= P-value; PP H4= posterior probability of H4 (full colocalization); Q= Q statistic; Q_df= Q degrees of freedom; SD= standard deviation; SE= standard error; SNP PP= posterior probability explained by SNP; 95% CI= 95% confidence interval

Supplementary Table S5: 95% credible sets for genomic regions with evidence for causal effects on kidney stone risk and colocalization

|  | Lead SNP from mineral metabolism genome wide association study in UK Biobank | | | | Colocalization analyses with kidney stone disease and 95% credible set | | | |
| --- | --- | --- | --- | --- | --- | --- | --- | --- |
| Trait | SNP | Gene | CHR | POS | N SNPs | PP H4 | Candidate causal SNP | PP |
| Adjusted calcium | rs11127048 | *GCKR* | 2 | 27752463 | 1750 | 0.87 | rs1260326 | 0.67 |
|  |  |  |  |  |  |  | rs780094 | 0.11 |
|  |  |  |  |  |  |  | rs780093 | 0.11 |
|  |  |  |  |  |  |  | rs4665972 | 0.07 |
| Adjusted calcium | rs11126078 | *MEIS1-AS2* | 2 | 66656402 | 3140 | 0.83 | rs2049019 | 0.21 |
|  |  |  |  |  |  |  | rs1519102 | 0.15 |
|  |  |  |  |  |  |  | rs7604427 | 0.10 |
|  |  |  |  |  |  |  | rs1519104 | 0.10 |
|  |  |  |  |  |  |  | rs6546230 | 0.09 |
|  |  |  |  |  |  |  | rs11126078 | 0.09 |
|  |  |  |  |  |  |  | rs756292 | 0.08 |
|  |  |  |  |  |  |  | rs13033745 | 0.07 |
|  |  |  |  |  |  |  | rs10183293 | 0.06 |
|  |  |  |  |  |  |  | rs6546232 | 0.04 |
| Adjusted calcium | rs2675966 | *C2orf82* | 2 | 233735543 | 3600 | 0.96 | rs11891546 | 1.00 |
| Adjusted calcium | rs838705 | *DGKD* | 2 | 234273242 | 3400 | 1.00 | rs838717 | 1.00 |
| Adjusted calcium | rs838717 | *DGKD* | 2 | 234296444 | 3360 | 1.00 | rs838717 | 1.00 |
| Adjusted calcium | rs13153019 | *SLC34A1* | 5 | 176782218 | 2390 | 0.8 | rs10051765 | 0.55 |
|  |  |  |  |  |  |  | rs13153019 | 0.13 |
|  |  |  |  |  |  |  | rs11748165 | 0.09 |
|  |  |  |  |  |  |  | rs56235845 | 0.08 |
|  |  |  |  |  |  |  | rs11741640 | 0.06 |
|  |  |  |  |  |  |  | rs4075958 | 0.05 |
| Adjusted calcium | rs2240736 | *BCAS3* | 17 | 59485393 | 1810 | 0.97 | rs9905274 | 0.48 |
|  |  |  |  |  |  |  | rs11657044 | 0.40 |
|  |  |  |  |  |  |  | rs9895661 | 0.09 |
| Adjusted calcium | rs209961 | *CYP24A1* | 20 | 52715154 | 3630 | 0.98 | rs6127099 | 0.98 |
| Adjusted calcium | rs2585442 | *CYP24A1* | 20 | 52737123 | 3670 | 0.98 | rs6127099 | 0.98 |
| Adjusted calcium | rs117268564 | *CYP24A1* | 20 | 52738434 | 3670 | 0.98 | rs6127099 | 0.98 |
| Adjusted calcium | rs35194449 | *CYP24A1* | 20 | 52742047 | 3670 | 0.98 | rs6127099 | 0.98 |
| Adjusted calcium | rs2762943 | *CYP24A1* | 20 | 52790786 | 3750 | 0.98 | rs6127099 | 0.98 |
| Phosphate | rs838718 | *DGKD* | 2 | 234296650 | 3370 | 0.99 | rs838718 | 0.48 |
|  |  |  |  |  |  |  | rs838717 | 0.47 |
|  |  |  |  |  |  |  | rs706848 | 0.04 |
| Phosphate | rs12152922 | *SLC34A1* | 5 | 176645599 | 2580 | 1.00 | rs10051765 | 0.82 |
|  |  |  |  |  |  |  | rs56235845 | 0.18 |
| Phosphate | rs10051765 | *SLC34A1* | 5 | 176799992 | 2360 | 1.00 | rs10051765 | 0.82 |
|  |  |  |  |  |  |  | rs56235845 | 0.18 |
| Phosphate | rs6127099 | *CYP24A1* | 20 | 52731402 | 3660 | 1.00 | rs35870583 | 0.65 |
|  |  |  |  |  |  |  | rs17216707 | 0.29 |
|  |  |  |  |  |  |  | rs6127099 | 0.06 |

CHR= chromosome; POS= position based on NCBI Genome Build 37 (hg19); N SNPs= number of SNPs; PP H4= posterior probability of H4 (full colocalization); SNP PP= posterior probability explained by SNP

Supplementary Table S6: Variants predicted to increase risk of kidney stones via effects on serum mineral metabolism

| Lead variant from mineral metabolism GWAS in UK Biobank | Candidate Gene | CHR | POS | Traits colocalizing in HyPrColoc | PP | Regional probability | Candidate causal variant | SNP PP |
| --- | --- | --- | --- | --- | --- | --- | --- | --- |
| rs56252443 | *GCKR* | 2 | 27465881 | KSD, albumin-adjusted serum calcium, serum phosphate, serum PTH | 0.99 | 0.99 | rs1260326 | 1.00 |
| rs11127048 | *GCKR* | 2 | 27752463 |  |  |  |  |  |
| rs838705 | *DGKD* | 2 | 234273242 | KSD, albumin-adjusted serum calcium, serum phosphate, serum PTH | 1.00 | 1.00 | rs838717 | 1.00 |
| rs838717 | *DGKD* | 2 | 234296444 |  |  |  |  |  |
| rs838718 | *DGKD* | 2 | 234296650 |  |  |  |  |  |
| rs13153019 | *SLC34A1* | 5 | 176782218 | KSD, albumin-adjusted serum calcium, serum phosphate, serum PTH | 1.00 | 1.00 | rs10051765 | 0.97 |
| rs12152922 | *SLC34A1* | 5 | 176645599 |  |  |  |  |  |
| rs10051765 | *SLC34A1* | 5 | 176799992 |  |  |  |  |  |
| rs209961 | *CYP24A1* | 20 | 52715154 | KSD, albumin-adjusted serum calcium, serum phosphate, serum PTH | 1.00 | 1.00 | rs6127099 | 1.00 |
| rs2585442 | *CYP24A1* | 20 | 52737123 |  |  |  |  |  |
| rs117268564 | *CYP24A1* | 20 | 52738434 |  |  |  |  |  |
| rs35194449 | *CYP24A1* | 20 | 52742047 |  |  |  |  |  |
| rs2762943 | *CYP24A1* | 20 | 52790786 |  |  |  |  |  |
| rs6127099 | *CYP24A1* | 20 | 52731402 |  |  |  |  |  |

CHR= chromosome; POS= position based on NCBI Genome Build 37 (hg19); PP= posterior probability of full colocalization; SNP PP= posterior probability explained by single nucleotide polymorphism (SNP); KSD= kidney stone disease; PTH=parathyroid hormone.

Supplementary Table S7: Associations of variants predicted to cause kidney stones with kidney stone disease and mineral metabolism traits in the UK Biobank

| Candidate causal variant | CHR | POS | Gene | Colocalizing traits | EA | NEA | EAF | KSD | | Adjusted calcium | | Phosphate | |
| --- | --- | --- | --- | --- | --- | --- | --- | --- | --- | --- | --- | --- | --- |
|  |  |  |  |  |  |  |  | OR  (95% CI) | P | Beta  (SE) | P | Beta  (SE) | P |
| rs838717 | 2 | 234296444 | *DGKD* | KSD, adjusted calcium, phosphate, PTH | G | A | 0.43 | 1.10  (1.07-1.13) | 2.60x10-11 | 0.05  (0.002) | 1.40x10-94 | -0.03  (0.002) | 7.2x10-36 |
| rs10051765 | 5 | 176799992 | *SLC34A1* | KSD, adjusted calcium, phosphate, PTH | C | T | 0.33 | 1.16  (1.13-1.19) | 1.30x10-24 | 0.03  (0.003) | 5.30x10-22 | -0.05  (0.003) | 9.80x10-90 |
| rs6127099 | 20 | 52731402 | *CYP24A1* | KSD, adjusted calcium, phosphate, PTH | A | T | 0.72 | 1.14  (1.10-1.17) | 1.60x10-16 | 0.06  (0.003) | 7.6x10-97 | 0.02  (0.003) | 1.40x10-13 |

CHR= chromosome; POS= position based on NCBI Genome Build 37 (hg19); EA= effect allele; NEA=non effect allele; EAF= effect allele frequency; KSD= kidney stone disease; P= p-value;

Supplementary Table S8: Associations of variants predicted to cause kidney stones with kidney stone disease and mineral metabolism traits in DiscovEHR

| **Variant** | **Effect allele** | **Gene** | **Kidney stone disease prevalence** | | **Serum calcium** | | **Serum phosphate** | |
| --- | --- | --- | --- | --- | --- | --- | --- | --- |
|  |  |  | OR | p | Beta | p | Beta | p |
| rs838717 | G | *DGKD* | 1.06 (1.04-1.09) | <0.0001 | 0.02 | <0.0001 | -0.02 | <0.0001 |
| rs10051765 | C | *SLC34A1* | 1.10 (1.08-1.12) | <0.0001 | 0.01 | <0.0001 | -0.04 | <0.0001 |
| rs6127099 | A | *CYP24A1* | 1.05 (1.03-1.08) | <0.0001 | 0.02 | <0.0001 | 0.01 | 0.02 |
| All | *G, A, C* | *DGKD, CYP24A1, SLC34A1* | 1.07 (1.06-1.09) | <0.0001 | 0.01 | <0.0001 | - | - |

Mean serum calcium and phosphate are adjusted for kidney stone disease case status. Odds ratio (OR) and beta reflect increased odds of kidney stone disease or change in biochemical measurement, respectively, with addition of one allele. Associations of combinations of *DGKD-, CYP24A1,* and *SLC34A1* risk alleles (All) were not assessed (-) for serum phosphate due to a lack of directional concordance.

Supplementary Table S9: Kidney stone population attributable risk and population attributable fraction in the UK Biobank

|  | **Risk allele carrier** | **No risk allele** | **Total kidney stone prevalence** | **Kidney stone prevalence non-carriers** | **Kidney stone prevalence carriers** | **PAR** | **PAF**  **(%)** |
| --- | --- | --- | --- | --- | --- | --- | --- |
| *DGKD* | | | 0.023 | 0.021 | 0.024 | 2.20x10^-3^ | 9.43 |
| rs838717 | GG/GA | AA |  |  |  |  |  |
| Case | 7994 | 3365 |  |  |  |  |  |
| No stones | 319985 | 156051 |  |  |  |  |  |
| *SLC34A1* | | |  |  |  |  |  |
| rs10051765 | CC/CT | TT | 0.023 | 0.022 | 0.025 | 1.63 x10^-3^ | 7.01 |
| Case | 6709 | 4650 |  |  |  |  |  |
| No stones | 266130 | 209906 |  |  |  |  |  |
| *CYP24A1* | | |  |  |  |  |  |
| rs6127099 | AA/AT | TT | 0.023 | 0.020 | 0.024 | 3.56 x10^-3^ | 15.29 |
| Case | 10610 | 749 |  |  |  |  |  |
| No stones | 438844 | 37192 |  |  |  |  |  |
| *DGKD, CYP24A1, SLC34A1* | | |  |  |  |  |  |
| All | 1+ risk allele | No risk alleles | 0.023 | 0.019 | 0.023 | 4.42x10^-3^ | 18.95 |
| Case | 11256 | 103 |  |  |  |  |  |
| No stones | 470686 | 5350 |  |  |  |  |  |

PAR= Population attributable risk (calculated as prevalence in total population- prevalence in individuals without risk allele); PAF= Population attributable fraction (calculated as (prevalence in total population- prevalence in individuals without risk allele)/ prevalence in total population)

Supplementary Table S10: Kidney stone population attributable risk and population attributable fraction in DiscovEHR.

|  | **Risk allele carrier** | **No risk allele** | **Total kidney stone prevalence** | **Kidney stone prevalence non-carriers** | **Kidney stone prevalence carriers** | **PAR** | **PAF**  **(%)** |
| --- | --- | --- | --- | --- | --- | --- | --- |
| *DGKD* | | | 0.113 | 0.106 | 0.116 | 6.12x10^-3^ | 5.44 |
| rs838717 | GG/GA | AA |  |  |  |  |  |
| Case | 13121 | 5839 |  |  |  |  |  |
| No stones | 100427 | 49000 |  |  |  |  |  |
| *SLC34A1* | | |  |  |  |  |  |
| rs10051765 | CC/CT | TT | 0.113 | 0.107 | 0.117 | 6.08x10^-3^ | 5.40 |
| Case | 11357 | 7603 |  |  |  |  |  |
| No stones | 85653 | 63774 |  |  |  |  |  |
| *CYP24A1* | | |  |  |  |  |  |
| rs6127099, | AA/AT | TT | 0.113 | 0.108 | 0.113 | 4.23x10^-3^ | 3.76 |
| Case | 17454 | 1506 |  |  |  |  |  |
| No stones | 137036 | 12391 |  |  |  |  |  |
| *DGKD, CYP24A1, SLC34A1* | | |  |  |  |  |  |
| All | 1+ risk allele | No risk alleles | 0.113 | 0.100 | 0.113 | 1.26x10^-2^ | 11.23 |
| Case | 18767 | 193 |  |  |  |  |  |
| No stones | 147689 | 1738 |  |  |  |  |  |

PAR= Population attributable risk (calculated as prevalence in total population- prevalence in individuals without risk allele); PAF= Population attributable fraction (calculated as (prevalence in total population- prevalence in individuals without risk allele)/ prevalence in total population)

Supplementary Table S11. Drug target Mendelian randomization; effects of modulating mineral metabolism traits on odds of kidney stone disease

| **Candidate gene target** | | | | | **Mendelian randomization with kidney stone disease** | | | | | | | | | | |
| --- | --- | --- | --- | --- | --- | --- | --- | --- | --- | --- | --- | --- | --- | --- | --- |
|  |  |  |  |  | **Sensitivity analysis** | **Instrumental variables** | | | **IVW Estimate** | | **Intercept** | | **Heterogeneity** | | |
| **Study** | **Gene** | **CHR** | **Start** | **End** | **Independence r2 threshold** | **Trait** | **N SNPs** | **Mean R2**  **(SD)** | **OR (95% CI)** | **FDR-P** | **Beta**  **(SE)** | **P** | **Q** | **Q_df** | **P** |
| CaSR-signaling | | | | | | | | | | | | | | | |
| UK Biobank | *DGKD* | 2 | 234263153 | 234380750 | 0.1 | Decreasing serum calcium* | 9 | 2.70x10-4  (3.1x10-4) | 0.26  (0.17-0.38) | 9.52x10^-11^ | -0.04  (0.02) | 0.06 | 10.76 | 8 | 0.22 |
| FinnGen | *DGKD* | 2 | 234263153 | 234380750 | 0.1 | Decreasing serum calcium* | 11 | 2.70x10-4  (3.1x10-4) | 0.21  (0.13-0.34) | 1.99x10^-10^ | -0.03  (0.02) | 0.35 | 15.88 | 10 | 0.10 |
| UK Biobank | *CASR* | 3 | 121902530 | 122005342 | 0.1 | Decreasing serum calcium* | 43 | 6.90x10-4  (9.2x10-4) | 0.69  (0.59-0.82) | 5.55x10^-5^ | 0  (0.01) | 0.94 | 110.52 | 42 | 0.00 |
| UK Biobank | *CASR* | 3 | 121902530 | 122005342 | 0.01 | Decreasing serum calcium* | 13 | 8.40x10-4  (1.3x10-3) | 0.67  (0.48-0.93) | 0.033 | 0.04  (0.04) | 0.28 | 41.42 | 12 | 0.00 |
| FinnGen | *CASR* | 3 | 121902530 | 122005342 | 0.1 | Decreasing serum calcium* | 35 | 6.90x10-4  (9.2x10-4) | 0.84  (0.70-0.99) | 0.04 | 2.31x10-3  (0.01) | 0.82 | 53.36 | 34 | 0.00 |
| FinnGen | *CASR* | 3 | 121902530 | 122005342 | 0.01 | Decreasing serum calcium* | 10 | 8.40x10-4  (1.3x10-3) | 0.75  (0.53-1.08) | 0.12 | 0.01  (0.04) | 0.74 | 26.73 | 9 | 0.00 |
| Phosphate metabolism | | | | | | | | | | | | | | | |
| UK Biobank | *SLC34A1* | 5 | 176806236 | 176825849 | 0.1 | Increasing serum phosphate | 8 | 2.80x10-4  (3.5x10-4) | 0.07  (0.04-0.1) | 3.47x10^-41^ | 0.02  (0.01) | 0.15 | 4.38 | 7 | 0.74 |
| FinnGen | *SLC34A1* | 5 | 176806236 | 176825849 | 0.1 | Increasing serum phosphate | 7 | 2.80x10-4  (3.5x10-4) | 0.07  (0.05-0.12) | 1.41x10^-29^ | 2.14x10-3  (0.02) | 0.90 | 2.16 | 6 | 0.90 |
| Vitamin D metabolism | | | | | | | | | | | | | | | |
| UK Biobank | *CYP24A1* | 20 | 52769988 | 52790512 | 0.1 | Decreasing serum calcium* | 17 | 3.10x10-4  (3.0x10-4) | 0.12  (0.09-0.15) | 1.03x10^-60^ | 0  (0.01) | 0.98 | 15.74 | 16 | 0.47 |
| UK Biobank | *CYP24A1* | 20 | 52769988 | 52790512 | 0.01 | Decreasing serum calcium* | 5 | 5.20x10-4  (5.1x10-4) | 0.12  (0.08-0.19) | 9.14x10^-18^ | -0.03  (0.03) | 0.45 | 6.29 | 4 | 0.18 |
| FinnGen | *CYP24A1* | 20 | 52769988 | 52790512 | 0.1 | Decreasing serum calcium* | 18 | 3.10x10-4  (3.0x10-4) | 0.09  (0.06-0.12) | 9.05x10^-50^ | -0.04  (0.01) | 0.01 | 22.22 | 18 | 0.18 |
| FinnGen | *CYP24A1* | 20 | 52769988 | 52790512 | 0.01 | Decreasing serum calcium* | 5 | 5.20x10-4  (5.1x10-4) | 0.07  (0.05-0.12) | 7.45x10^-27^ | -0.02  (0.03) | 0.62 | 5.49 | 4 | 0.24 |

* Albumin-adjusted serum calcium concentration; CHR= chromosome; End= end position; FDR-P= P-value adjusted for 5% false discovery rate; N SNPs= number of SNPs; P -value; Q= Q statistic; Q_df= Q degrees of freedom; SD= standard deviation; SE=standard error; Start= start position

Supplementary Table S12: Rare predicted deleterious *DGKD* missense variants in 100kGP participants with kidney stone disease

| Variable | Normal Range | Proband 1 | Proband 2 | Proband 3 |
| --- | --- | --- | --- | --- |
| *DGKD* variant | - | His190Gln | Ile221Asn | Arg1181Trp |
| *DGKD* variant ID |  | 2:233434885:C:G | 2:233435893:T:A | 2:233468539:C:T |
| Sex | - | Female | Female | Male |
| Stone analysis | - | UK | UK | Calcium oxalate 70%, calcium phosphate 30% |
| Adjusted serum calcium (mmol/l) | 2.10-2.50 | UK | UK | 2.45 |
| Serum phosphate (mmol/l) | 0.70-1.40 | UK | UK | 1.14 |
| 24-hour urinary calcium excretion (mmol) | >5 | UK | UK | 8.9 |
| 24-hour urinary pH | - | UK | UK | 6.86 |
| 24-hour urinary phosphate excretion (mmol) | <35 | UK | UK | 29.6 |
| 24-hour urinary oxalate excretion (umol) | <460 | UK | UK | 273 |
| 24-hour urinary citrate excretion (mmol) | >2.5 | UK | UK | 2.39 |
| 24-hour urinary magnesium excretion (mmol) | >3 | UK | UK | 5.2 |
| 24-hour urinary uric acid excretion (umol) | 200-430 males  140-360 females | UK | UK | 400 |
| Family history of kidney stones | - | UK | Yes | Yes |
| Additional phenotypes | - | Hyperparathyroidism | - | - |
| Allele frequency 100KGP* | - | 9.4x10^-5^ | 3.9x10^-5^ | 6.7x10^-4^ |
| Allele frequency DiscovEHR | - | 1.4x10^-4^ | - | 1.9x10^-3^ |
| Allele frequency Gnomad | - | 5.0x10^-5^ | - | 6.0x10^-4^ |
| CADD | - | 11 | 24 | 32 |
| SIFT | - | 0.02 | 0 | 0 |
| PolyPhen | - | 0.768 | 0.987 | 1 |

*Frequency in rare diseases HG38 cohort; UK unknown.

Supplementary Table S13: Rare *DGKD* missense variants associated with kidney stone disease in the DiscovEHR cohort

| *DGKD*  Variant | Variant ID | Controls n (%) | Stone cases n (%) | P value | CADD | SIFT | Polyphen | Allele frequency Gnomad | Allele frequency 100kGP* | Allele frequency DiscovEHR |
| --- | --- | --- | --- | --- | --- | --- | --- | --- | --- | --- |
| Ile91Val | 2:233390406:A:G | 14 (0.01) | 5 (0.03) | 0.049 | 21 | 0.01 | 0.102 | 3.5x10-5 | - | 1.4x10^-4^ |
| Thr319Ala | 2:233438249:A:G | 1 (<0.1) | 2 (0.01) | 0.033 | 20 | 0.26 | 0.00 | - | - | 2.2x10^-5^ |
| Val464Ile | 2:233446767:G:A | 25 (0.02) | 8 (0.06) | 0.023 | 22 | 0.03 | 0.003 | 1.1x10-4 | 7.1x10-5 | 2.4x10^-4^ |
| Arg900His | 2:233459761:G:A | 5 (<0.01) | 3 (0.02) | 0.048 | 27 | 0 | 0.954 | 2.8x10-5 | - | 5.9x10^-5^ |

*Frequency in rare diseases HG38 cohort.

5.	HapMap 3 - Wellcome Sanger Institute. https://www.sanger.ac.uk/resources/downloads/human/hapmap3.html.

6.	Altshuler, D. M. *et al.* International HapMap 3 Consortium: Integrating common and rare genetic variation in diverse human populations. *Nature* **467**, 52.

7.	Yang, J. *et al.* Conditional and joint multiple-SNP analysis of GWAS summary statistics identifies additional variants influencing complex traits. *Nat. Genet.* **44**, 369–375 (2012).

8.	Yang, J., Lee, S. H., Goddard, M. E. & Visscher, P. M. GCTA: a tool for genome-wide complex trait analysis. *Am. J. Hum. Genet.* **88**, 76–82 (2011).

9.	Weir, B. S. *Genetic Data Analysis. Methods for Discrete Population Genetic Data.* (Sinauer Associates, Inc. Publishers, 1990).
